# Lymphodepletion mitigates anti-CAR immunity in pediatric and young adult patients with recurrent or refractory brain tumors: clinical trial results

**DOI:** 10.64898/2026.08.27.26361261

**Authors:** Leo D. Wang, Angela M. Taravella Oill, Silke E. Lindner, Tracey Stiller, M. Suzette Blanchard, Rishika V. Mudunuri, Melody Wu, Jonathan C. Hibbard, Colt Egelston, Sean M. Sepulveda, Lance Peter, Julie L. Kilpatrick, Jennifer Stratman, Evan D. Mee, Daniel G. Chen, Giacomo Oliveira, Margarita Muñoz, Jamie Wagner, Ally Mardiroos Dolatabadi, Monica Nisis, Jennifer K. Shepphird, Gabriela Sanchez, Angela Burmayan, Kiana Langeliers, Jinny Paul, Yolanda Villasenor, Jack Wadden, Tiffany Adam, Daniel de la Nava, Heini M. Natri, Cheryl Oliver-Cervantes, Lisa Feldman, Shilpa Shahani, Maryam Aftabizadeh, Stephen J. Forman, Julie Ressler, Leonidas Arvanitis, Katie M. Campbell, Jennifer A. Cotter, Sarah A. Richman, Massimo D’Apuzzo, Benita Tamrazi, Behnam Badie, Catherine J. Wu, Jay A. Read, Santhosh A. Upadhyaya, Carl Koschmann, Tom B. Davidson, Nicholas E. Banovich, Christine E. Brown

## Abstract

Outcomes for high-grade pediatric brain tumor patients remain poor, but there is optimism that chimeric antigen receptor (CAR) T cell therapy can improve prognosis. We present the results from a phase I clinical trial of IL13BBζ-CAR T cells infused weekly into the cerebral ventricles in pediatric and young adult patients with recurrent or refractory brain tumors. The trial met its primary objectives of feasibility, safety, and tolerability, with one dose-limiting toxicity. 8 of 16 patients evaluable for response experienced radiographic size decreases consistent with biologic activity and with an anti-tumor response. Two patients met protocol criteria for response. Median survival for patients receiving lymphodepletion was 20.5 months from diagnosis and 6.9 months from treatment for patients with midline glioma, and 187 months from diagnosis and 7.5 months from treatment for patients with ependymoma. Importantly, patients who did not receive lymphodepletion developed anti-CAR humoral and cellular immune responses detectable in the CSF and peripheral blood, whereas patients receiving lymphodepletion had no evidence of CSF anti-CAR immunity. Taken together, these findings demonstrate the safety, tolerability, and biological activity of locoregionally-delivered IL13BBζ-CAR T cells for children and young adults with CNS tumors. Moreover, we show that anti-CAR immune responses arise in patients not receiving lymphodepletion, but not in the CSF of patients receiving systemic lymphodepletion. Further investigation of adoptive cellular therapies combined with immunosuppression is warranted in this patient population. ClinicalTrials.gov registration: NCT04510051.

## Introduction

Outcomes for patients with recurrent or aggressive brain tumors remain poor, and standard treatments are associated with significant morbidity^1–7^. New therapies are urgently needed, and chimeric antigen receptor (CAR) T cell therapy has shown promise in both pediatric^8–13^ and adult^14–22^ brain tumors. Ongoing investigations in the field evaluate the effects of target selection, CAR construction, route of delivery, and lymphodepletion on CAR efficacy; current pediatric brain tumor CAR T trials target GD2, B7-H3, HER2, and IL13Rα2 among other antigens^14,23–26^. An important open question in these trials is whether lymphodepletion should be incorporated, as is required for hematologic malignancy CAR T therapies. Comparing outcomes and correlative analyses across these trials is critical for improving these therapies as quickly as possible.

Our group recently published the results of a large phase I clinical trial testing the efficacy of IL13Rα2-targeting CAR T cells in adults with glioblastoma multiforme^22^. This trial showed encouraging evidence of clinical activity in a large cohort of heavily pretreated adult patients, with a few patients experiencing remarkable responses^15,22^. Our chimeric antigen receptor is directed against the high-affinity IL13 receptor, IL13Rα2, and is composed entirely of human derived components. Unlike most CAR designs that employ single-chain variable fragments for antigen recognition, this construct uses a modified high affinity cytokine ligand as its binding domain, bearing a single E13Y substitution to minimize off-target interactions^23,27^. The codon optimized CAR is fused to a human IgG4 Fc spacer containing L235E and N297Q mutations to abrogate Fc receptor mediated interactions, followed by a CD4-derived transmembrane domain, a 4-1BB-derived costimulatory domain, and a human CD3ζ cytoplasmic signaling domain. This CAR has been extensively evaluated in preclinical and adult clinical studies^14,15,19,23,28^. Our group has previously shown that CAR T cells are more effective when manufactured from less mature initial T cell populations^29,30^, and more effective when administered locoregionally, both in preclinical animal models^23,31^ and in clinical trial settings^15^. For this reason, our trials incorporated these manufacturing advancements and included locoregional delivery into the cerebrospinal fluid. Finally, preclinical^14,24^ and early clinical evidence indicates that systemic lymphodepletion improves the efficacy of locoregionally-delivered CAR T cells^8,32^. To test the impact of this intervention on CAR T cell persistence, host immune dynamics, and the development of anti-CAR immune responses, we incorporated lymphodepletion into this trial.

Here, we report results from the first two cohorts of pediatric and young adult patients treated with intraventricular IL13BBζ-CAR T cells on this phase I clinical trial, testing for the first time the combination of IL13BBζ-CAR T cells with lymphodepletion (NCT04510051). Patients ≥4 and ≤24 years old (≤21 years old at initial diagnosis) with recurrent or refractory primary neuromalignancies were eligible to enroll provided their tumor tissue was confirmed to express IL13Rα2, and they had progressed after standard therapy. We demonstrate safety of repeated weekly intracranial infusions of IL13BBζ-CAR T cells, with or without preceding lymphodepletion, as well as evidence of biological activity and immune activation in the CNS. Moreover, we identify anti-CAR immune responses emerging in patients on therapy, which are significantly suppressed by lymphodepletion.

## Results

### Trial design and patient characteristics

The primary objective of this ongoing study is to assess the safety and feasibility of repeated intraventricular (ICV) delivery of IL13Rα2-directed CAR T cells with or without prior systemic lymphodepleting chemotherapy in children and young adults with IL13Rα2^+^ recurrent or refractory brain tumors (NCT04510051).

Given the demonstrated safety of IL13Rα2-targeting CAR T cells in adults with brain tumors^19^, the documented superiority of locoregional delivery of these cells in animal models^23,31^, and the preclinical and early clinical evidence supporting lymphodepletion for locoregionally-delivered solid tumor-targeted CAR T cells^8,32^, our trial design incorporates lymphodepletion before intraventricular IL13BBζ-CAR T cell therapy for pediatric and young adult patients with documented IL13Rα2^+^ recurrent or refractory brain tumors.

Patients received weekly intraventricular IL13BBζ-CAR T cell infusions, with the first infusion at a dose of 10 × 10^6^ CAR^+^ T cells and subsequent infusions at 50 × 10^6^ CAR^+^ T cells (**Fig. 1a,b**). The first three patients on the trial (cohort 1) did not receive lymphodepletion. The subsequent 15 patients (cohort 2) received four doses of fludarabine (30 mg/m^2^/day) and two doses of cyclophosphamide (500 mg/m^2^/day), followed by a rest day before the first IL13BBζ-CAR T cell infusion. After the first four infusions, encompassing the dose limiting toxicity (DLT) evaluation period, patients had the option to continue regular intraventricular infusions and to undertake other cancer-directed therapies (**Fig. S1**). During active therapy, patients underwent sampling of cerebrospinal fluid (CSF) and peripheral blood (PB) immediately before and 1–2 days after each CAR T infusion.

**Fig. 1.**
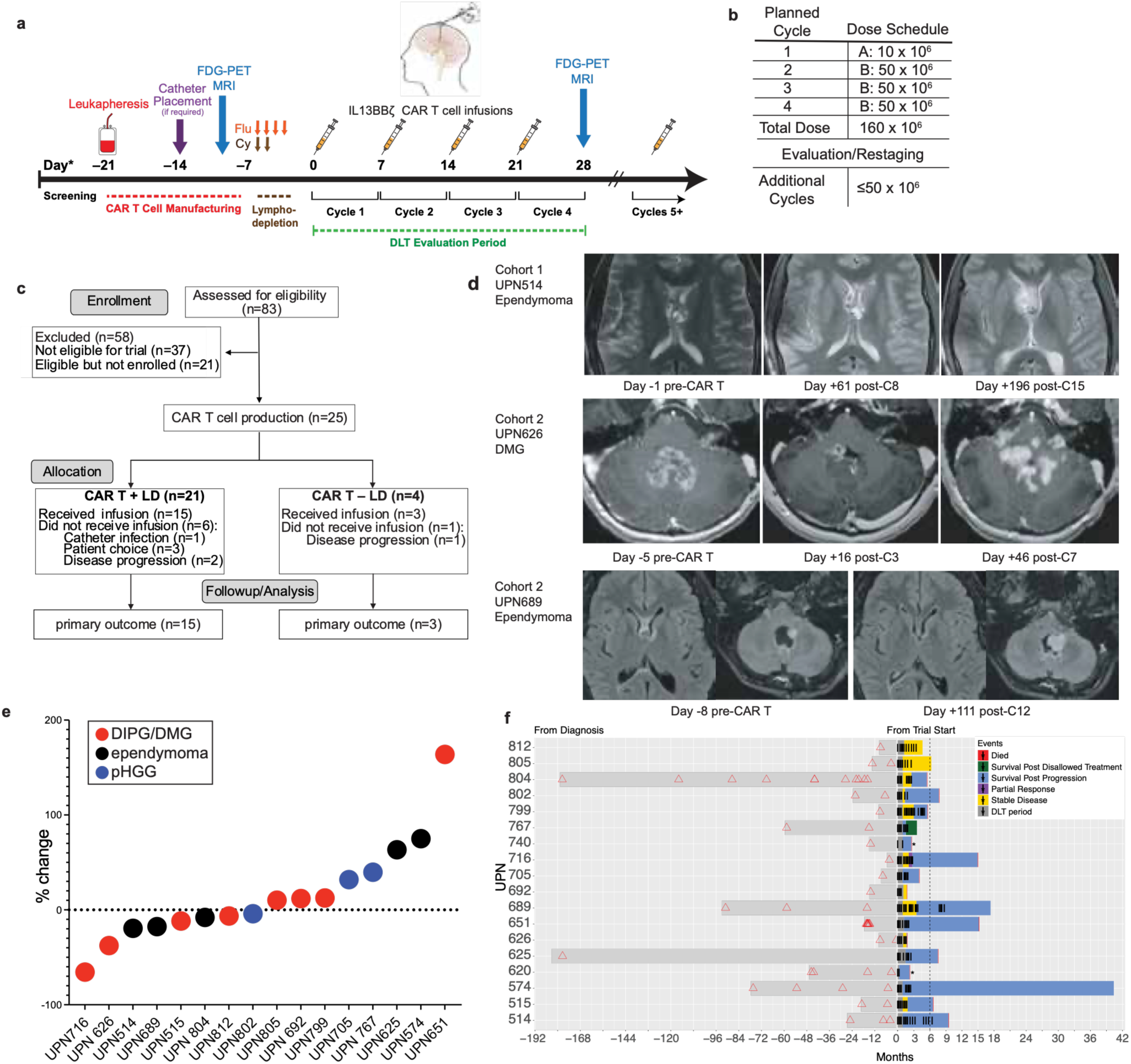
Trial schema and clinical activity of IL13BBζ CAR T cell therapy. **a.** Treatment schema for patients enrolled in cohorts 1 and 2. **b.** Dose schedule for IL13BBζ CAR T cell administration. **c**. CONSORT flow diagram showing participant enrolment and allocation for both cohorts. **d.** Representative MRI images from participants in cohorts 1 and 2 obtained before CAR T cell infusion and at the indicated post-treatment time points. **e.** Waterfall plot showing the largest percentage decrease from baseline in measurable tumor burden for each individual patient, as assessed by MRI. Colors denote disease type: ependymoma (black), pediatric high-grade glioma (pHGG; blue), and diffuse intrinsic pontine or midline glioma (DIPG/DMG; red). **f.** Swimmer plot summarizing clinical outcomes for 18 patients. Each bar represents an individual participant from diagnosis to trial enrollment (grey) and during treatment (colored). Red triangles denote radiation therapy, black lines indicate intraventricular (ICV) CAR T cell infusion cycles, and asterisks mark patients who did not complete the required four treatment cycles.

### Patient characteristics and treatment

Eligibility criteria included age ≥4yo and ≤24yo at the time of treatment and age ≤21yo at the time of diagnosis, histopathologic diagnosis of a primary brain tumor confirmed to have progressed or recurred after the completion of standard therapy, performance score ≥60, dexamethasone dose ≤0.1mg/kg/day up to 6mg, protocol-defined washout from other therapies, and protocol-defined laboratory and organ function criteria. Immunohistochemical screening for IL13Rα2 was required to document an H-Score ≥ 50 for eligibility.

As of December 31^st^, 2025, 83 patients had been screened by immunohistochemistry (IHC), of whom 46 (55%) had qualifying H scores (**Table S1**). Of these, 25 participants completed leukapheresis and product manufacture successfully; there was one failed product (**Fig. 1c, Table S2**). Eighteen patients received CAR T therapy. The median age of our patient population was 19 yrs (range: 11-25 yrs), reflective of our enrollment of patients with recurrent/progressive disease. The seven patients who did not receive treatment opted for other therapies (n = 2) or progressed during manufacture and did not meet criteria for CAR T infusion (n = 5, representing 20% of patients completing leukapheresis). Patients on this trial were assigned unique patient identifiers that were not known to anyone outside the research group. No patient underwent repeat leukapheresis. Baseline characteristics are reported in **Table 1**.

**Table 1.** Patient characteristics. Cohort 1 comprised 3 patients who did not receive systemic lymphodepletion before treatment. Cohort 2 comprised 15 patients who received lymphodepletion. Two patients in cohort 2 were not evaluable for response; UPN620 developed an intercurrent infection unrelated to therapy and UPN740 experienced phrenic nerve paralysis attributed to TIAN, qualifying as a DLT. Otherwise, all patients completed at least 4 and as many as 18 infusions (median: 8). Concomitant dexamethasone use was permitted up to 0.1mg/kg/day (max: 6mg). Best radiographic responses and vital status as of 12/2025 are noted. Ages are represented as quartile range of enrolled patients (ages 11-24).

| Table 1. Patient characteristics |  |  |  |  |  |  |  |
| --- | --- | --- | --- | --- | --- | --- | --- |
| Research Subject | Age/ Sex 1st infusion | Diagnosis | Cycles received | KPS/ Lansky | Steroids/ Bevacizumab | Best Response# | Vital Status |
| Cohort 1 |  |  |  |  |  |  |  |
| UPN514 | 18-21/M | Posterior fossa group A (PFA) ependymoma, grade 3 | 15 | 90 | Dexamethasone | PD | Deceased |
| UPN515 | 14-17/F | DIPG, H3K27-altered, grade 4 | 8 | 70 | Dexamethasone | SD | Deceased |
| UPN574 | 22-24/F | PFA ependymoma, grade 3 | 9 | 100 |  | PD | Alive |
| Cohort 2 |  |  |  |  |  |  |  |
| UPN620 | 22-24/M | High grade astrocytoma, H3G34R, grade 4 | 2 | 50 |  | Unevaluable | Deceased |
| UPN625 | 14-17/M | Posterior fossa group A (PFA) ependymoma grade 3 | 8 | 60 | Dexamethasone | PD | Deceased |
| UPN626 | 14-17/M | DMG, H3K27-altered, grade 4 | 7 | 60 | Dexamethasone | SD | Deceased |
| UPN651 | 22-24/M | DMG, H3K27-altered, BRAFV600E, grade 4 | 8 | 80 |  | PD | Deceased |
| UPN692 | 18-21/M | DMG, H3K27-altered, grade 4 | 4 | 70 | Dexamethasone | SD | Deceased |
| UPN689 | 22-24/M | Ependymoma, NOS, grade 3 | 18 | 80 |  | SD | Alive |
| UPN705 | 18-21/M | pHGG, H3-wildtype, IDH-wildtype, grade 4 | 8 | 60 | Dexamethasone | PD | Deceased |
| UPN716 | 18-21/M | DMG, H3 K27-altered, grade 4 | 12 | 90 | Dordaviprone | PR | Deceased |
| UPN740 | 14-17/M | DMG, H3K27-altered, grade 4 | 2 | 60 | Dexamethasone | Unevaluable | Deceased |
| UPN767 | 22-24/F | High-grade glioma, consistent with pleomorphic Xanthoastrocytoma, grade 3 | 8 | 90 | Bevacizumab | PseudoPD | Alive |
| UPN799 | 14-17/M | DMG, H3K27-altered, grade 4 | 13 | 70 | Dexamethasone | SD | Deceased |
| UPN802 | 22-24/M | pHGG, H3-wildtype, IDH-wildtype, CMMRD, grade 4 | 6 | 70 | Dexamethasone | SD | Alive |
| UPN804 | 22-24/M | Ependymoma, grade 3 | 9 | 90 | Dexamethasone | SD | Deceased |
| UPN805 | 18-21/M | DMG, H3K27-altered, grade 4 | 7 | 60 |  | SD | Alive |
| UPN812 | 11-13/M | DMG, H3K27-altered, grade 4 | 9 | 60 |  | SD | Alive |

### Safety

All patients who received one or more CAR T infusions are included in the AE summary (**Table S3**). Overall, CAR T therapy with or without lymphodepletion was well-tolerated in these patients, with one DLT observed (phrenic nerve paralysis leading to respiratory compromise) during the 28-day observation period encompassing the first four infusions. One patient did not complete the DLT period due to an intercurrent infection. The most common toxicities attributed to CAR T at the level of possible or probable were CRS, nausea, vomiting, fatigue, headache, and fever (**Table S3**). These toxicities were generally mild and self-limited (mostly grade 1 and 2), with headache developing metronomically generally 24-36 hours after each infusion (median 1 day, range 1-2 days) before resolving with minimal intervention. More severe toxicities attributed to CAR T as possible or above included grade 3 gait disturbance, somnolence, hypoxia, headache, seizure, increased liver enzymes, and grade 3-4 cytopenias in patients receiving lymphodepletion. Patients were permitted to use steroids and bevacizumab as anti-inflammatory therapy if needed (**Table 1; Fig. S1**). Because IL13Rα2 is also expressed on testis^33^, we amended our protocol in September 2022 to collect testosterone data on patients. Some subsequent male patients were noted to have low pre-treatment testosterone levels, but otherwise patient testosterone levels fluctuated throughout therapy (**Fig. S2**). In these patients, an association between CAR T therapy and testosterone level was not observed. Overall, CAR T therapy with or without lymphodepletion seems safe and well-tolerated.

### Treatment outcomes

Patients in cohorts 1 and 2 either had recurrent or refractory ependymoma or recurrent/refractory pediatric high-grade glioma (pHGG), including brainstem and midline glioma (DIPG/DMG). Patients received a median of 8 (range: 2-19) CAR T cell infusions (**Table 1**). Because the natural histories of these diseases are quite different, we present survival statistics by disease group. At the time of analysis, 11 of the 16 evaluable patients were deceased. Some patients chose to initiate other tumor-targeted therapy after the DLT period (**Fig. S1**); these patients were excluded from subsequent survival analysis. Median survival from time of treatment for ependymoma patients receiving lymphodepletion was 7.5 months (95%CI 5.4-not estimable); median survival from diagnosis was 187.4 months (95%CI 184.1-not estimable). For DIPG/DMG patients receiving lymphodepletion, median survival from treatment was 6.9 months (95%CI 1.71-not estimable) and median survival from diagnosis was 20.5 months (95%CI 16.5-not estimable). For other lymphodepleted pHGG patients, median survival from treatment was 5.2 months (95% 3.98-not estimable) and median survival from diagnosis was 29.9 months (95% 12.6-not estimable)(**Table 2**).

**Table 2.** Median survival. Of the patients who received lymphodepletion, 7 had DIPG/DMG, 3 had ependymoma, and 3 had pHGG. 3 patients did not receive lymphodepletion; one had DIPG and two had ependymoma. Median survival from time of treatment (top) and from diagnosis (bottom) is noted in months, with corresponding 95% confidence intervals. Because patients remained alive at the data cutoff, the 95% CI upper limit was not calculable.

| Table 2. Median survival |  |  |  |  |  |
| --- | --- | --- | --- | --- | --- |
| from treatment |  |  |  |  |  |
|  | patients | deaths | median (mo) | 0.95LCL | 0.95UCL |
| DMG/DIPG | 7 | 5 | 6.9 | 1.71 | NA |
| Ependymoma | 3 | 2 | 7.49 | 5.42 | NA |
| pHGG | 3 | 2 | 5.16 | 3.98 | NA |
| No Lymphodepletion | 3 | 2 | 9.46 | 6.57 | NA |
| from diagnosis |  |  |  |  |  |
|  | patients | deaths | median (mo) | 0.95LCL | 0.95UCL |
| DMG/DIPG | 7 | 5 | 20.5 | 16.5 | NA |
| Ependymoma | 3 | 2 | 187.4 | 184.1 | NA |
| pHGG | 3 | 2 | 29.9 | 12.6 | NA |
| No Lymphodepletion | 3 | 2 | 36.1 | 25.8 | NA |

Among the patients in this report who completed the DLT evaluation period (n = 16) the best individual responses by MRI using RAPNO criteria were one partial response (PR; 6.25%), one minor response (minR; 6.25%); 10 stable diseases (SD; 62.5%), and four progressive diseases (PD; 25%)(**Table 3**). Improvements in all cases were transient, lasting no longer than 12 cycles. In the initial cohort, receiving CAR T cells alone, two of three evaluable participants achieved stable disease (**Table 3**). Although only two patients achieved radiographic response per protocol, half (8 of 16) had mixed radiographic responses, with at least some tumor masses decreasing in size with treatment (**Figure 1d-f, Table S4**). Taken together, these results indicate that IL13BBζ-CAR T cells can confer radiographic benefit to pediatric and young adult patients with IL13Rα2-expressing brain tumors.

**Table 3.** Best protocol responses. Of patients evaluable for response, 6/7 patients with DIPG/DMG experienced stable disease or better radiographic responses by RAPNO criteria. One patient experienced a minor response, and one experienced a partial response. Of the 3 ependymoma patients, all of whom were evaluable, 2 had stable disease; both of these had mixed responses with some lesions shrinking and others not (see Fig. 1d). Of the 3 evaluable pHGG patients, one achieved stable disease.

| Table 3. Response - N (%) |  |  |  |  |
| --- | --- | --- | --- | --- |
| Characteristic | DMG/DIPG | Ependymoma | pHGG | Without Lymphodepletion |
|  | N = 8<sup>1</sup> | N = 3<sup>1</sup> | N = 4<sup>1</sup> | N = 3<sup>1</sup> |
| Best Response |  |  |  |  |
| Minor Response/Partial Response (minR/PR) | 2 (29%) | 0 (0%) | 0 (0%) | 0 (0%) |
| Stable Disease (SD) | 4 (57%) | 2 (67%) | 1 (33%) | 2 (67%) |
| Progressive Disease (PD) | 1 (14%) | 1 (33%) | 2 (67%) | 1 (33%) |
| Not Evaluable | 1 | 0 | 1 | 0 |
| n (%) |  |  |  |  |

### Correlative studies

#### Peripheral blood and CSF immune landscapes are affected by lymphodepletion

In both the peripheral blood and CSF, we quantified lymphocyte counts across patients and cycles using clinical cell counts (**Fig. 2a-b**; see Methods). Immediately following lymphodepletion (cycle 1 day 0) we observed significant lymphocyte reductions in the peripheral blood compared to patients who did not receive lymphodepletion (p = 0.01 Mann-Whitney U from cycle 1 day 0; **Fig 2b**). However, by 7 weeks post-lymphodepletion (treatment cycle 8) lymphocytes normalized to the level of the non-lymphodepleted patients (**Fig. S3**). In the CSF we observed lower lymphocyte counts compared to peripheral blood, irrespective of lymphodepletion status. While patients receiving lymphodepletion tended to have lower CSF lymphocyte counts in the first weeks post treatment, this pattern was not as clear as in peripheral blood (**Fig. 2a**). To more deeply characterize and contrast the immune dynamics with and without lymphodepletion, we performed scRNA-seq on PBMCs, CSF, and engineered product samples from a subset of patients and timepoints (**Table S5**).

**Fig. 2.**
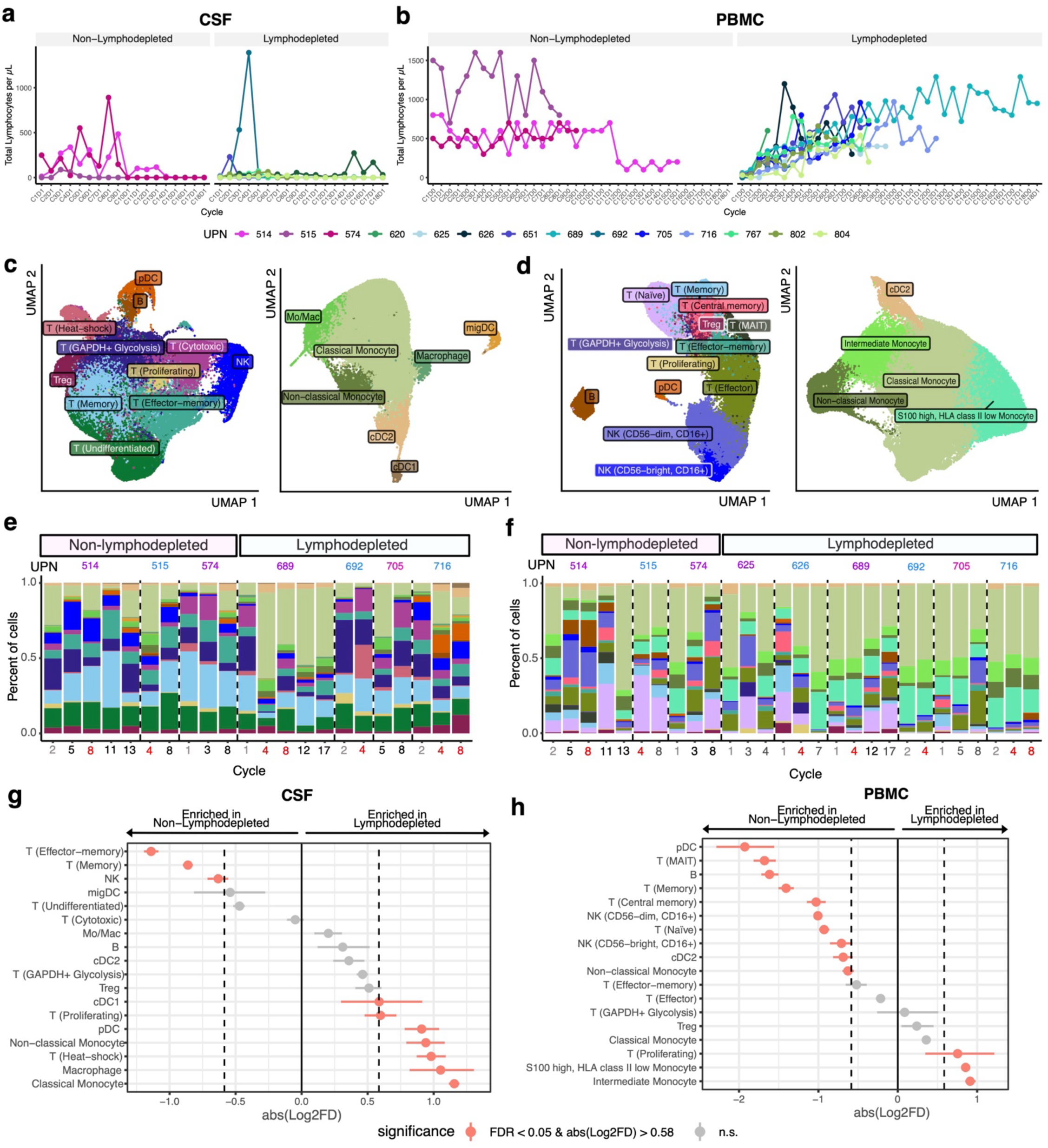
Single-cell landscape of CSF and PB immune cells over the course of CAR T cell therapy. Total lymphocyte counts per μL calculated from clinical counts for **a**) CSF and **b**) PBMC stratified by patient lymphodepletion status. Colors in **a-b** correspond to patients (UPN). UMAPs of **c**) 86,047 cells from CSF lymphoid lineage (left) and 28,753 cells from CSF myeloid lineage (right), and **d**) 41,034 cells from PBMC lymphoid lineage (left) and 99,362 cells from PBMC myeloid lineage (right). **e,f**) Stacked bar plot of the proportion of cell types from CSF and PBMCs, respectively, colored according the UMAPs in **c**) and **d**). Colors represent cell types. Bars are labeled UPN (top) and cycle treatment cycle number (bottom); grey indicates baseline cycles, black indicates cycles that roughly correspond to nonresponse periods; red indicates response periods; boxes labeled lymphodepleted correspond to patients that received lymphodepletion prior to treatment and boxes labeled non-lymphodepleted correspond to patients that did not have lymphodepletion. **g-h**) Point range plots comparing the proportion of cell types between patients that received lymphodepletion and patients that did not in **(g)** CSF and **(h)** PBMC. Red points in **g-h** represent cell types with a significant proportional difference between categories and the red horizontal lines represent 95% CIs. Dashed vertical lines represent absolute log2 fold difference (abs(Log2FD)) of 0.58. Significant proportional differences between categories were ones with both FDR <0.05 and abs(Log2FD) of 0.58. Solid vertical line represents abs(Log2FD) of 0. abs(Log2FD) greater than 0 represents cell types at a higher proportion in patients that received lymphodepletion compared to those who did not.

We recovered in total 114,800 cells across 18 immune cell types and states in CSF, 140,396 cells across 18 immune cell types and states in PBMCs, and 10,885 cells across 5 T cell states in engineered product (**Fig. 2c,d, Fig. S4-S8**). When stratified by patient and cycle, we observed that cell type proportions fluctuate over the course of therapy, although no specific patterns emerged over successive cycles of therapy (**Fig. 2e,f**). In particular, others have reported the infiltration of proinflammatory myeloid populations into the CSF concurrent with intraventricular CAR T cell therapy^34^; this was not a consistent finding in our patients. However, we observed compositional shifts in both CSF and peripheral blood of patients who received lymphodepletion (**Fig. 2g,h, Table S6-7**). Of note, in the CSF, we observed a significant enrichment of effector memory T cells in patients who did not receive lymphodepletion as compared to those who did (FDR < 0.05 and abs(Log2FD) > 0.58; **Fig. 2g**, **Fig. S9**, **Table S6**). More specifically, when comparing early treatment cycles to late treatment cycles (see Methods), we observed an increase in effector memory T cells in both lymphodepleted and non-lymphodepleted, but significantly higher accumulation in the non-lymphodepleted patients (**Fig. S10**). For the scRNA-seq analyses we chose timepoints that coincided with prescheduled MRI studies, acknowledging that imaging correlates imperfectly with the kinetics of response, but selecting timepoints where radiographic changes were most apparent (**Fig. 1d,e**). Interestingly, we also observed a decrease in the proportion of effector memory T cells during periods of radiographic response compared to periods of non-response – although these changes were not statistically significant (**Fig. S11**). Taken together, these data suggest lymphodepletion is altering the immune landscape in both the peripheral blood and CSF and at least some of the changes (e.g. reduction of effector memory T cells) are also observed in periods of radiographic response.

Finally, we assessed the degree to which expression programs were coordinated across PBMCs and CSF. Analyzing each cell type independently, we tested for associations between gene expression levels in CSF and PBMCs, e.g., if higher expression of a given gene in peripheral blood T cells corresponded with higher expression of that gene in CSF T cells. We observed 4,261 gene-cell type pairs out of a total of 67,587 gene-cell type pairs with a significant relationship between these two compartments (**Fig. S12, Table S8**). Of these, a majority (2,408) of significant genes were observed in T cells, followed by monocytes (984), NK cells (685), cDC2s (149), and B cells (35)(**Fig. S12**) – suggesting for some immune compartments peripheral blood may be a good proxy for immunophenotyping the CSF.

#### Single-cell analysis reveals clonal expansion of CAR^-^ T cells in CSF but not peripheral blood

Using a modification of the standard scRNAseq workflow, we selectively enriched RNA fragments mapping to the CAR construct, enabling us to categorize T cells as CAR^+^ or CAR^−^ (See Methods). Both CAR^+^ and CAR^−^ T cells were detected in CSF and PBMC, with the majority being CAR^−^ (**Figure 3a, Table S9, S10**). CAR^+^ T cells were generally more abundant in CSF than in PBMC across all patients and all timepoints (**Table S9, S10**). However, absolute cell count analysis of the CSF from the flow cytometry data showed that some patients had substantially higher numbers of CAR^+^ T cells in their CSF than others; all of these patients received lymphodepletion (**Fig. 3b**). CAR^−^ T cells had higher proportions of Tregs, effector-memory and undifferentiated phenotypes, while CAR^+^ T cells had a higher proportion of heat-shock, cytotoxic, GAPDH+ glycolysis and proliferating phenotypes in CSF (FDR < 0.05 and abs(Log2FD) > 0.58; **Fig. 3c, Table S11**).

**Fig. 3.**
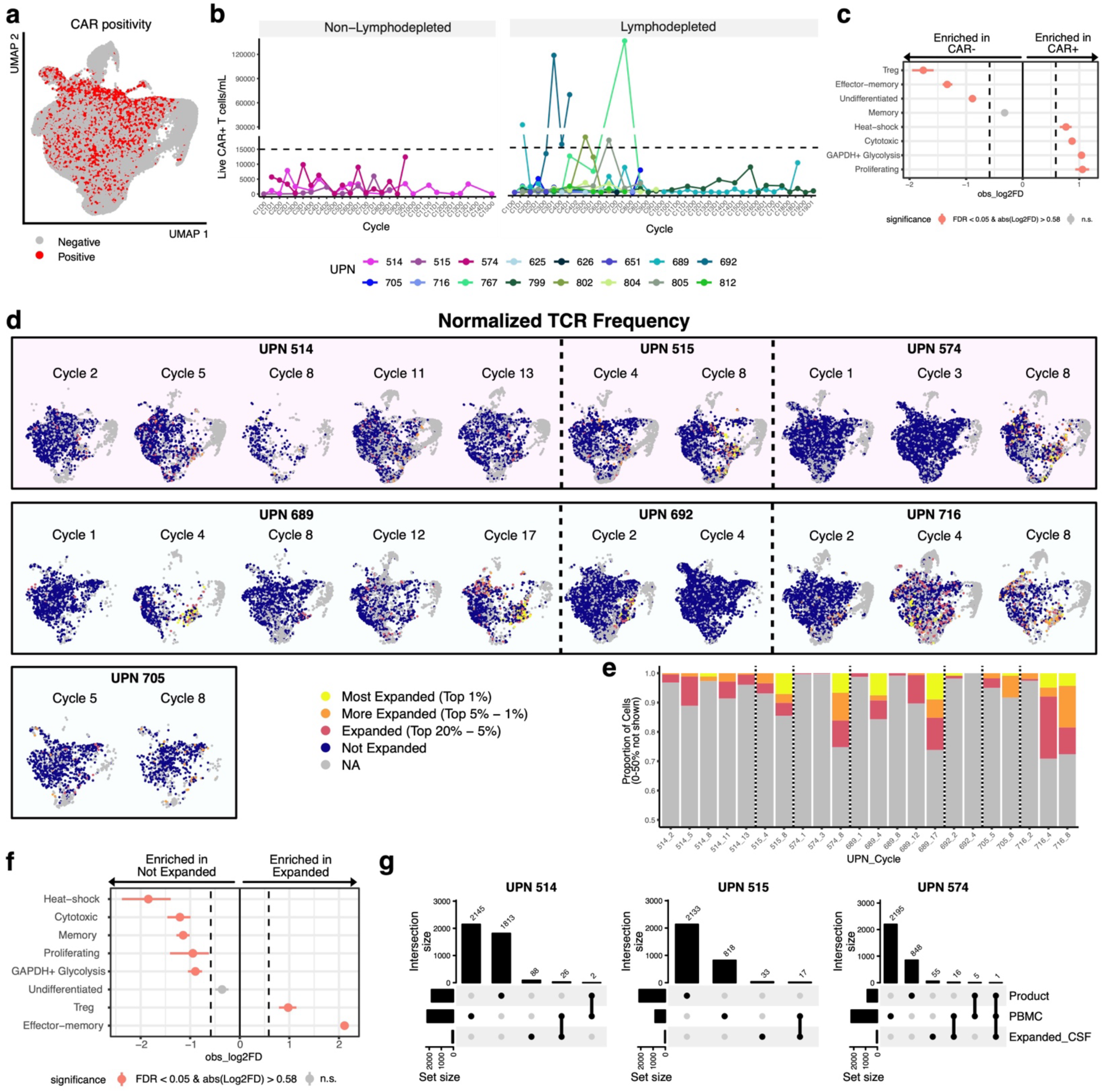
CAR-T and clonotype dynamics of T cells over the course of therapy. **a,** UMAP of CSF lymphoid cells colored by CAR positivity. Red dots are CAR+ T cells, grey are all other cells. **b.** Live CAR⁺ T cells per mL of total cells in CSF from non-LD (top) and LD (bottom) participants. Measurements were obtained at time points defined by the study protocol, including prior to CAR T-cell infusion and 24 hours post-infusion. **c.** Point range plots comparing the proportion of T cell states in CSF between CAR+ T cells and CAR-T cells. **d.** UMAP of CSF lymphoid cells split by patient (UPN) and cycle. Cells are color-coded by normalized TCR frequency category; yellow denotes most expanded (top 1% of all detected TCRs) and orange denotes more expanded (top 1–5% of all detected TCRs) at each timepoint. **e.** Bar graph demonstrates that representation of expanded TCRs increases over time for all patients. **f.** Point range plot comparing the proportion of T cell states in CSF between cells with expanded (Top 20%) and unexpanded TCRs. **g.** Upset plots comparing expanded TCRs in CSF to all TCRs in the product and PBMC for each patient. Red points in **c** and **f** represent T cell subsets/states with a significant proportional difference between categories; red lines represent 95% CI. Dashed vertical lines represent absolute log2 fold difference (abs(Log2FD)) of 0.58. Significant proportional differences between categories were ones with both FDR <0.05 and abs(Log2FD) of 0.58. Solid vertical line represents abs(Log2FD) of 0. abs(Log2FD) greater than 0 represents cell types at a higher proportion in CAR+ T cells compared to CAR-T cells or among cells with an expanded TCR compared to ones that did not have an expanded TCR.

To understand the clonal dynamics of T cells during the course of CAR T therapy we also generated scTCR-seq data for these patients. Unique TCR sequences can be used as cellular barcodes to trace clonally related cells in the scRNA-seq data as all cells bearing the same TCR share a common progenitor. This analysis revealed the presence of multiple clonally related T cells in the CSF. Intriguingly, clonally expanded T cells in the CSF were generally CAR-negative (**Fig. 3d, Fig. S13**). Across cycle timepoints there was an increase in the proportion of the most expanded TCR clonotypes in all patients; this was true for all patients when comparing early and late timepoints although the magnitude of increase was variable (**Fig. 3e**). This suggests continued expansion over time. Among cells with expanded TCRs, we observed a higher proportion of effector memory cells and Tregs compared to cells that were not expanded (FDR < 0.05 and abs(Log2FD) > 0.58; **Fig. 3f, Table S12**). Interestingly, with the exception of one TCR clonotype in one patient (UPN574), TCRs found to be expanded in the CSF were not observed in the product and were infrequently observed in PBMCs (**Fig. 3g**). Importantly, TCRs found to be expanded in the CSF that were also identified in product and PBMC were not expanded in those compartments. Overall, our characterization of the T cell state landscape showed that endogenous CAR^−^ T cell clones were increasingly prevalent in the CSF over the course of CAR T cell therapy, and that these expanded T cells are highly enriched for effector memory phenotype. This suggests that CAR T cell therapy may entrain and/or expand endogenous activated effector T cells in the CSF.

### Identification of anti-CAR immune responses in patients not receiving lymphodepletion

Based on the observation that patients who received lymphodepletion often had higher absolute numbers of CAR T cells in their CSF than patients who did not, despite an identical weekly dosing protocol (**Fig. 3b**), we hypothesized that patients who did not receive lymphodepletion developed anti-CAR immune responses that restrained CAR T numbers. Thus, we screened patient peripheral blood plasma and CSF for anti-CAR antibodies. While no patient had preexisting anti-CAR antibodies, all three non-lymphodepleted patients developed anti-CAR antibodies detectable in peripheral blood and CSF, emerging at Cycle 5 or 6 of therapy. In contrast, patients who received lymphodepletion did not develop anti-CAR antibodies detectable in CSF, even after 18 infusion cycles. Two lymphodepleted patients developed anti-CAR antibodies in the serum with delayed kinetics (Cycles 7 and 11)(**Fig. 4a, Table S13**). To define the specificity of antibodies raised against the CAR construct, we screened CAR-reactive patient serum against a distinct, HER2-targeting, CAR construct^35^. This construct is comprised of a HER2-targeting scFv joined to an IgG4(EQ)-derived linker region, CD8 transmembrane, and 4-1BB costimulatory domain, and CD3ζ cytoplasmic tail. Extracellularly, the sole difference between the CARs is in the antigen-binding domain. We found that serum from UPN514 and UPN515 recognized the HER2 and IL13 CARs with the same intensity and temporal kinetics, strongly suggesting that the antibodies recognized the IgG4-Fc linker region common to both CARs; this linker contains mutations at L235E and N297Q to reduce FcR interactions. In contrast, antisera from UPN574, UPN651, and UPN812 did not react against the HER2 CAR, suggesting that they recognize the antigen-binding mutein that is specific to the IL13 CAR (**Fig. 4b**).

**Fig. 4.**
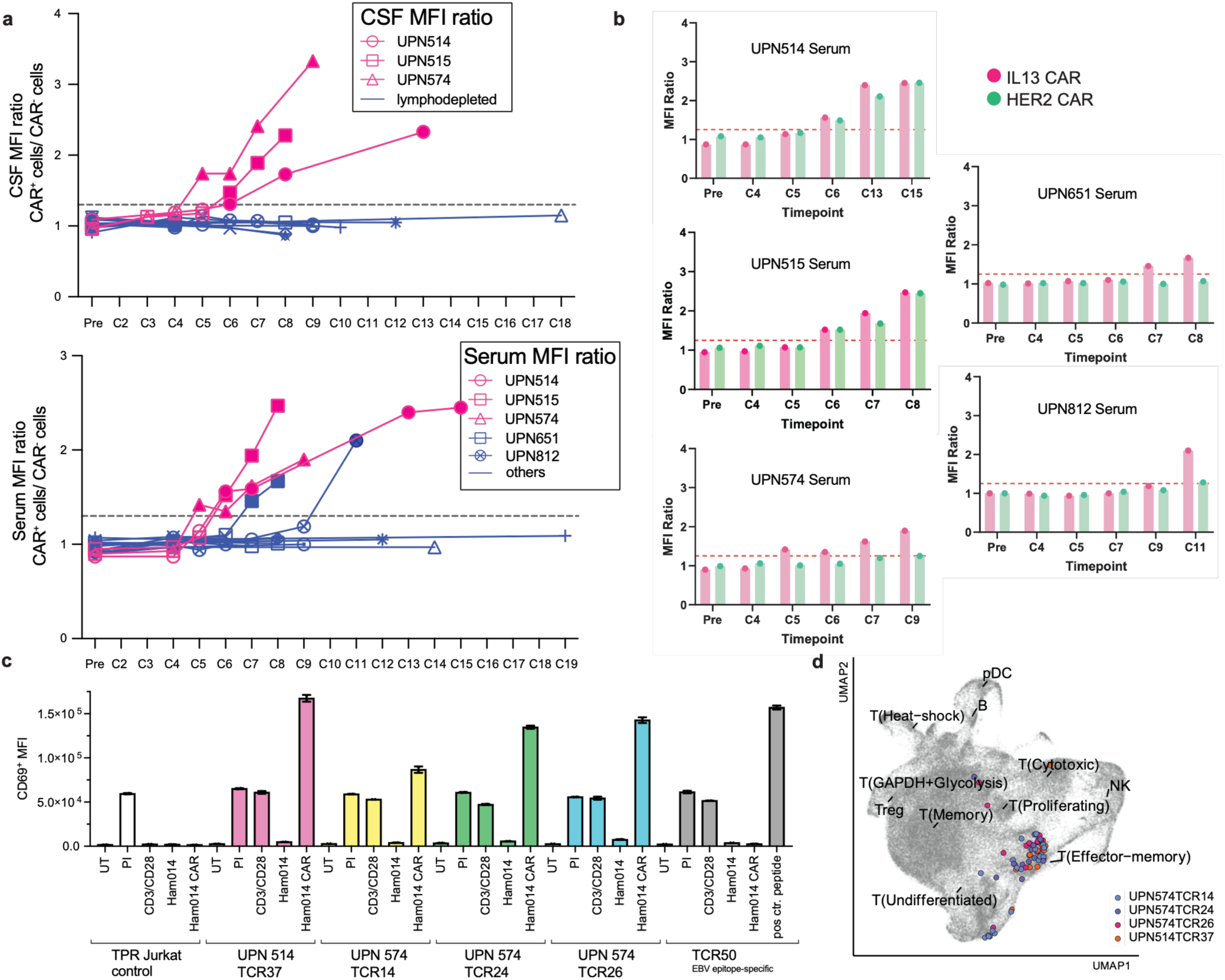
Detection of anti-CAR immune responses in patients receiving CAR T cell therapy. Anti-CAR antibodies were quantified by sandwich flow cytometry across serum and CSF samples and reported as the ratio of mean fluorescence intensities (MFIs) of staining on CAR-expressing cells compared to CAR-negative cels. **a.** In CSF (top), anti-CAR antibodies (filled symbols) were detected in non-lymphodepleted patients (magenta) at cycle 5 (UPN574) or 6 (UPN514, 515). No anti-CAR antibodies were detected in the CSF of patients who received lymphodepletion (blue). In peripheral blood serum (bottom), anti-CAR antibodies were again detected in non-lymphodepleted patients, at the identical cycle timepoints (5 and 6). Anti-CAR antibodies were detected in 2 lymphodepleted patients, but at cycles 7 and 11. Anti-CAR antibody assays were done in triplicate and repeated 2-3 times for each patient and timepoint; positive results were confirmed with an additional replicate. **b.** In patients who developed anti-CAR antibodies, serum was screened against HER2-CAR expressing Jurkat cells, which share only the IgG4-based linked domain extracellularly. UPN514 and UPN515 appeared to recognize both CARs with equivalent kinetics, suggesting overlap in specificity and therefore IgG4 binding. UPN574, UPN651, and UPN812 did not recognize the HER2-CAR, suggesting that they recognize the IL13 mutein. **c**. TCR activation was assessed by CD69 upregulation on Jurkat T cells, as measured by flow cytometry. TPR Jurkat cells, which do not express TCR, responded only to PMA-ionomycin (PI). All other cell lines expressed a TCR and responded both to PI and to CD3/28 antibody stimulation. TCR50, which recognizes an EBV LMP2 epitope, was used as a positive control for MHC expression on Ham014 APCs. Reporter cells expressing TCR37 (UPN514) or TCR14, TCR24, or TCR26 (UPN574) upregulated CD69 when cocultured with APCs expressing the IL13BBζ CAR, but not when cocultured with CAR^-^ APCs. Bar graph is representative of 3 independent experiments, each done in technical duplicate. **d**, UMAP showing CAR-recognizing T cells, with clonotypes color-coded by patient and TCR number. Most CAR-specific T cells are in the effector memory cluster.

We further hypothesized that anti-CAR humoral immune responses would be accompanied by clonal expansion of CAR-reactive effector memory T cells. To test this, expanded T cell receptor (TCR) sequences identified in the CSF of patients who did not receive lymphodepletion were cloned and expressed in Jurkat reporter cells, which were subsequently cocultured with partially HLA-matched target cells expressing the CAR construct. Four CAR-reactive TCRs were identified in two patients (TCR37 in UPN514; and TCRs 14, 24, and 26 in UPN574). TCR37 was detected after cycle 11 of therapy, whereas TCRs 14, 24, and 26 were detected following cycle 8 of therapy (**Fig. 4c**). By mapping the CAR-reactive TCR clonotypes back to scRNAseq data, we confirmed that the majority exhibited an effector memory phenotype **(Fig. 4d**). Taken together, these data clearly demonstrate that patients receiving intraventricular IL13BBζ-CAR T cells can develop anti-CAR B and T cell immunity, and that this anti-CAR immunity is significantly mitigated by systemic lymphodepletion.

### Virus-specific endogenous T cells also expand in the CSF during CAR T cell therapy

Many clonally-expanded T cells in the CSF of patients not receiving lymphodepletion were not CAR-reactive, and we were interested in the specificities of these T cells given their transcriptional and immunophenotypic identities. Intriguingly, we identified three TCRs in UPN514 (TCR12, TCR13 and TCR42) specific for Epstein Barr virus-derived peptides. These TCRs were predominantly associated with a granzyme K-expressing effector memory T cell phenotype, as has been previously described^36^ (**Fig. S14b,c**). While we were not able to identify active EBV epitope expression in residual tumor sample from these patients, it is well-documented that anti-EBV T cell responses can lead to neuroimmunity^37–40^. Thus, these findings raise the provocative possibility that antiviral immune responses sparked by CAR T therapy can be leveraged to improve antitumor immune control.

### CSF cell-free DNA sequencing and variant allele fraction analysis

Circulating tumor DNA (ctDNA) sequencing has been validated in the CSF as a technique for determining tumor burden in a variety of brain tumors^41,42^, and tumor-informed ctDNA sequencing has been used in other pediatric neurooncology clinical trials to track treatment response and predict progression^43^. We hypothesized that ctDNA could be useful in evaluating tumor response at early timepoints in patients receiving CAR T cell therapy, especially as immunotherapy-mediated tumor swelling can render radiographic changes difficult to interpret. We therefore submitted pre-treatment and Cycle 4 Day 0 (pre-Cycle 4) CSF for tumor-informed ctDNA sequencing for three patients who did not receive lymphodepletion and three patients who did (**Fig. S15**). In many patients, changes in ctDNA variant allele frequency (VAF) paralleled radiographic changes: UPN515 had a decrease in tumor size concomitant with a decrease in VAF, and UPN 705 had increases in both tumor size and VAF. UPN514 had stable disease over the first 4 cycles of therapy, and then experienced a mixed decrease in tumor size between cycles 4 and 8; VAF analysis in this patient shows an increase in one mutation but not in another, perhaps consistent with a mixed response. Similarly, UPN716 experienced a mixed radiographic response that was also a partial response by RAPNO criteria; VAF analysis suggests that this partial response may partly be attributable to tumor evolution. UPN574 and UPN692 both had decreases in ctDNA VAF, but both also experienced tumor progression by the end of Cycle 4. Taken together, these findings suggest that ctDNA analysis may be a useful adjunct for tracking tumor response, as well as a potential role for multitarget measurement in evaluating tumor clonal evolution.

### Single-cell analysis of post-treatment tumor samples

Two patients, UPN574 and UPN625, underwent tumor resection after CAR T therapy. UPN574 had a resection one week after Cycle 9 of CAR T therapy, and UPN625 underwent resection one month after Cycle 8 of CAR T therapy. Both patients had PFA ependymoma, and neither experienced a radiographic response during treatment. UPN574 did not undergo lymphodepletion, whereas UPN625 did. Both of their resected tumors had low levels of T cell infiltration and high levels of myeloid infiltration post-CAR therapy by immunohistochemistry, similar to pre-therapy; these features have been well described as a poor prognostic indicator associated with recurrence and with subgroup A ependymoma^44–46^. Both resected tumors also showed preserved expression of IL13Rα2 (data not shown). Notably, UPN574 demonstrated metronomic spikes in inflammatory cytokines in the CSF, indistinguishable from patients who experienced a radiographic mixed response (**Fig. S16**). Unfortunately, CSF samples from UPN625 were insufficient for single-cell analyses. For UPN574, however, clonal expansion of CAR^−^ effector memory T cells in the CSF was a prominent feature and indistinguishable from the kinetics observed in the CSF of UPN514 and UPN515 (**Fig. 3**). Resected tumors were submitted for single-cell sequencing analysis, and we recovered a total of 1,891 immune cells across 5 cell types from those two patients (**Fig. S17, S18**). All of the 233 intratumoral T cells observed were in an activated state and all observed T cells were CAR^−^ with various TCR frequencies (**Fig. S17a-c**). Some TCRs were observed to overlap with TCRs observed to be expanded in CSF (**Fig. 3g, Fig. S17d**). For UPN625, where we obtained scRNAseq for pre- and post-treatment tumors, we observed lower proportions of cDC2s, mast cells, and T cells and higher proportions of mo/mac cells in post-treatment tumor compared to pretreatment tumor (**Fig. S17e**).

Comparisons of overlap of TCRs among data types revealed enrichment of expanded CSF TCRs in tumor. For UPN574, we obtained scTCRseq only for post treatment tumor, finding expanded CSF TCRs were significantly enriched for tumor overlapping clones (Fisher’s exact test, p < 2.2×10^-16^, OR = 248). Of the 72 expanded clonotypes in CSF, 17 overlapped with tumor clonotypes, while out of 13,869 non-expanded CSF clonotypes, 17 overlapped with tumor (**Fig. S17d**). Taken together, these data suggest that endogenous CAR^−^ T cells can traffic between CSF and tumor, and that expanded endogenous T cells may do this more effectively than unexpanded T cells. CAR^+^ T cells were not detected in post-therapy tumors, which may be due to poor access to the tumor, poor persistence in the tumor, or both. Notably, CAR^+^ T cells were also not prominently detected in the CSF sample from UPN574 obtained closest to resection (Cycle 8, **Table S9**). These findings highlight the importance of evaluating both the CSF and the tumor itself, as some features of CSF may correlate with features in the intratumoral microenvironment, whereas others may not. Additionally, these data underscore the importance of local tumoral obstacles in limiting the potential efficacy of adoptive cellular therapies for solid tumors.

## Discussion

We present results from the first two cohorts of pediatric and young adult patients with recurrent or refractory high grade brain tumors treated intraventricularly with IL13BBζ-CAR T cells, with or without preceding lymphodepletion. We show the therapy to be safe and well-tolerated. Over half of patients experienced some radiographic, albeit transient, benefit. This radiographic mixed response, as well as median overall survival for patients on trial, is similar to other reports testing CAR T cell therapies in children and young adults with these diseases^12,13,34^.

This is also, to our knowledge, the first description of anti-CAR humoral and cellular immune responses in brain tumor patients. In all patients not receiving lymphodepletion, we discovered anti-CAR antibodies arising simultaneously in both the CSF and blood serum, emerging at the 5^th^ or 6^th^ cycle of therapy. CSF anti-CAR antibodies were not detected in any patient receiving lymphodepletion, even after as many as 18 infusions. Serum anti-CAR T cells were detected in 2/15 lymphodepleted patients, at cycles 7 and 11. Additionally, we identified clonally expanded, activated CD8^+^ T cells that specifically recognize CAR in an MHC-dependent manner, in patients who did not receive lymphodepletion. Moreover, although all patients received identical CAR T cell doses per cycle, we observed that patients who did not receive lymphodepletion had substantially lower CSF CAR T cell numbers than many patients who received lymphodepletion. These results are further supported by the scRNA-seq analyses which identified increased effector memory T cells in the CSF of patients that did not receive lymphodepletion. Furthermore, we found clonal expansion of T cells in the CSF was limited almost exclusively to CAR^−^ T cells, and that these cells were highly enriched for effector memory phenotype. At least some expanded TCRs were found to recognize CAR-specific peptides.

Our results strongly suggest that systemic lymphodepletion abrogates, or significantly mitigates, an anti-CAR immune response that may limit CAR T cell numbers in the CSF. More speculatively, the observation that anti-CAR antibodies can be found in both serum and CSF or in serum alone, but not in CSF alone, suggests a model in which anti-CAR immunity emerges first in the periphery and then expands into the CSF. This would presumably depend on adoptively transferred CAR T cells exiting the CSF and transiting to peripheral lymphoid organs to initiate a peripheral anti-CAR immune response that then finds its way back to the CSF. Given the well-described differences between the blood-CSF barrier and the blood-brain barrier, this proposed ‘boomerang’ mechanism may help to explain why intratumorally-delivered CAR T cells do not seem to engender anti-CAR immunity, whereas intraventricularly-delivered CAR T cells do^47^. Regardless of underlying mechanism, these findings clearly establish the importance of considering anti-CAR immunity in cellular therapies for pediatric brain tumors.

T cell clonal diversity in peripheral blood and tumor have been extensively studied (reviewed in ^48^ and elsewhere), but to our knowledge this is the first report of clonal expansion in the CSF of human patients receiving immunotherapy. The implications of TCR diversity and clonality in the context of immunotherapy are clearly dependent on tumor type, therapy, and other factors. However, the identification of a TCR specific for the CAR construct clearly indicates antagonistic cellular immunity in the absence of lymphodepletion. Conversely, the finding that some expanded TCRs recognize viral epitopes suggests that these might be leveraged therapeutically. Taken together, these findings may represent a first step in understanding how to use adoptive cellular therapies to promote endogenous antitumor immune responses.

Consistent with other studies, including our extensive experience with adult glioblastoma patients receiving IL13BBζ-CAR T cells^10,14^, and consistent with patients’ metronomic clinical symptoms, intraventricular delivery of CAR T cells was associated with post-infusion increases in CSF inflammatory cytokines such as IFNγ, IL-6, IL-8, CCL4, and CXCL10 (**Fig. S16**). These spikes were detected the day after each infusion and dissipated before the next CAR T infusion. Moreover, studies with IL13BBζ-CAR T cells in adult glioblastoma finds that levels of interferon pathway cytokines IFNγ, CXCL9, and CXCL10 are positively associated with clinical response^14^. Peripheral blood serum cytokine levels, in contrast, were much more stable both within and between cycles; this is also similar to what has been reported by other groups^8,10,19,23^. We did not see large differences in either CSF or serum cytokine profiles in patients receiving or not receiving lymphodepletion.

Our analysis of post-CAR T therapy tumors in two patients (UPN574 and 625) should not be overinterpreted. However, our data suggest that endogenous immune cells, particularly clonally expanded effector memory cells found in the CSF, may preferentially gain access to the tumor as compared to unexpanded T cells. CAR^+^ T cells were not recovered from post-therapy tumor, suggesting that they may have poor access to or persistence in the tumor microenvironment in nonresponding patients.

This initial report of pediatric and young adult patients treated with IL13Rα2-targeted CAR T cells indicates that intraventricular CAR T cell therapy is safe, well-tolerated, and active in young people with recurrent or refractory brain tumors, both with and without preceding lymphodepletion; these clinical results, including overall survival from time of diagnosis, are very similar to those reported by other groups. Additionally, this is, to our knowledge, the first report showing that systemic lymphodepletion can prevent or delay cellular and humoral immunosuppressive responses to CAR T therapy, resulting in increased CAR T cell numbers in the CSF over the course of therapy. Moreover, we report that intraventricular delivery of CAR T cells is associated with infiltration and clonal expansion of antiviral endogenous T cells in the CSF. These data have clear implications for further avenues of study in ongoing trials, and could only be generated by coupling longitudinal CSF, peripheral blood, and tumor sampling with single-cell techniques. Importantly, deep interrogation of post-therapy tissue will likely be critical for improving cellular therapies; every effort should be made to partner with patients and families to gain this precious knowledge. On the basis of these promising initial results, and in recognition of important observations from our colleagues at other institutions (reported in this issue), we have expanded this trial to treat newly diagnosed DIPG/DMG patients at our center and at two other sites. Enrollment and treatment of patients in this final cohort is ongoing.

## Data Availability

All data produced in the present study will be made available upon request after publication, subject to federal, state, and local regulations governing patient privacy

## Methods

### Oversight and informed consent

The clinical protocol and all amendments were approved by the City of Hope Institutional Review Board. Ongoing review is performed by a Data and Safety Monitoring Committee (DSMC) as well as a Cancer Protocol Review and Monitoring Committee (CPRMC). Patients and/or guardians provided written informed consent before tissue screening and leukapheresis, and a separate written informed consent before enrollment on the therapeutic portion of the trial. Minor assent was obtained when appropriate.

### Clinical trial design

This phase I study assesses the safety and feasibility of repeated intraventricular (ICV) delivery of IL13Rα2-directed CAR T cells after systemic lymphodepleting chemotherapy in children and young adults with IL13Rα2^+^ recurrent or refractory brain tumors (NCT04510051). Patients receive at least 4 weekly IL13Rα2-CAR T cell infusions, with the first infusion at a dose of 10 × 10^6^ CAR^+^ T cells and subsequent infusions at 50 × 10^6^ CAR^+^ T cells (**Fig. 1a**). To verify that CAR T cells administered alone were safe and well-tolerated, the first three patients on the trial did not receive lymphodepletion. Subsequent patients receive intravenous cyclophosphamide (500mg/m^2^ per day × 2 days, on days −5 and −4) and fludarabine (30mg/m^2^ per day × 4 days, on days −5 through −2). The first 3 participants on treatment plan 1, and the first 3 participants of each disease type on treatment plan 2 were staggered through 28 days. The primary objective of this study is to assess safety and feasibility of repeated intraventricular (ICV) delivery of IL13Rα2-directed CAR T cells. Secondary objectives include evaluating persistence and expansion of CAR T cells and endogenous immune cells in peripheral blood (PB) and cerebrospinal fluid (CSF), evaluating cytokine levels in PB and CSF, and evaluating potential efficacy and biologic response to treatment. Radiographic responses are evaluated using RAPNO criteria for ependymoma, DIPG/DMG, and HGG^49–52^ with the modification that images were assessed at 4 weeks and 8 weeks after the start of therapy, and approximately every 4 weeks thereafter. Lesions found to be larger at two consecutive imaging timepoints were deemed true progression.

Children and young adults under 21 years of age at time of initial diagnosis (ages 4–24 at the time of leukapheresis at City of Hope, and ages 1–24 at the University of Michigan and Children’s Hospital of Los Angeles) with histologically-confirmed malignant brain neoplasms are eligible, with radiographic evidence of recurrence or progression after the end of the initial conventional therapy, including radiation. Additional requirements include IL13Rα2 tumor expression by immunohistochemistry (IHC; H-score > 50) and a Karnofsky or Lansky performance score ≥ 60, except for loss of mobility due to disease.

The trial has been approved by the Institutional Review Board at City of Hope and is conducted under a Food and Drug Administration (FDA) Investigational New Drug application in accordance with International Council for Harmonization Good Clinical Practice guidelines. The first patient was treated on the trial December 14, 2020. Further details of the trial design are provided in the **Supplementary Appendix**.

### MRI acquisition and analysis

MRI of the brain and spine were acquired on a Siemens MAGNETOM Verio 3.0 Tesla scanner. Tumor foci were measured on axial T1 multi-planar reconstruction (MPR)-weighted images obtained after the administration of MultiHance (gadobenate dimeglumine) or Gadovist (gadobutrol). Response assessment was recorded using RAPNO criteria for pediatric brain tumors. For the calculation of contrast-enhancing tumor volumes, T2-weighted, T2-weighted Fluid Attenuated Inversion Recovery, T1-weighted pre- and post-contrast images were co-registered and resampled to 1 mm × 1 mm × 3 mm voxel sizes using BraTumIA software ^53^. The registered, non-skull-stripped images were then imported into ITK-SNAP (v3.8.0) for segmentation ^54^. Trained readers generated initial masks of the tumor volumes with final review and volume selection by neuroradiologists with over 10 years of experience in neuroradiology.

### IL13Ra2-targeted CAR design and manufacture

The design of our CAR construct has previously been described^14,23,28^. Briefly, the construct comprises a modified cytokine-based antigen binding domain coupled to an IgG4Fc-based hinge and linker region, followed by a CD4 transmembrane domain, 4-1BB costimulatory domain, and CD3ζ signaling domain. This construct was lentivirally transduced into a naïve/memory T cell pool, as previously documented^23,28^. For the 18 patients described here, product manufacturing characteristics included are detailed in **Table S2**.

### Patient sample processing

Clinical tests were run on CLIA-certified machines using industry standard protocols. CBCs and CSF cell counts were performed on a Sysmex XN-9100; differentials were done manually on the Cellavision platform. Chemistries were done by immunoassay using either a Beckman Coulter AU5800 or a Roche Cobas c503. Testosterone levels were run by immunoassay on a Roche Cobas Pro SBL e801 at City of Hope, and were done either by immunoassay or LC/MS-MS at University of Michigan. For research studies, peripheral blood was collected in vacutainer tubes with or without EDTA. EDTA-treated samples were processed immediately by Ficoll density-gradient centrifugation, and peripheral blood mononuclear cells (PBMCs) were cryopreserved in CryoStor CS5 at −80 °C before transfer to liquid nitrogen for long-term storage. Blood samples collected without EDTA were allowed to clot for 2–3 h at room temperature; serum was isolated by centrifugation, aliquoted into single-use volumes (100– 200 µl), and stored at −80 °C. Research CSF samples were centrifuged, and cell-free supernatants were aliquoted and stored at −80 °C. CSF cell pellets were resuspended in HBSS without calcium and magnesium (Corning CellGro) supplemented with 2% fetal calf serum (FCS) and sodium azide for immediate flow cytometric analysis. Remaining CSF cells were resuspended in CryoStor CS10, frozen at −80 °C, and subsequently transferred to liquid nitrogen for long-term storage. Tumor specimens were collected at Children’s Hospital Los Angeles (CHLA) and processed at City of Hope under an approved institutional review board protocol.

### Immunohistochemistry

Immunohistochemistry (IHC) was done on 5 µm-thick sections of formalin-fixed paraffin-embedded specimens placed on positively charged glass slides. The slides were loaded on a Ventana DISCOVERY ULTRA IHC automated stainer, where deparaffinization, rehydration, endogenous peroxidase activity inhibition and antigen retrieval (using TRIS buffer pH8) were first performed. The slides were then incubated with a monoclonal rabbit anti-human IL13Rα2 (E7U7B) (Cell Signaling Technology, diluted 1:100) for 32 minutes followed by incubation with agents from the OptiView DAB IHC Detection Kit. The stains were counterstained with hematoxylin and coverslipped. The stained slides were scanned using a NanoZoomer 2.0-HT digital slide scanner, a NanoZoomer S360 Digital Slide Scanner (Hamamatsu Corporation) or directly acquired on an Olympus BX46 transmitted light microscope with an SC-180 Olympus camera.

IL13Rα2 immunoreactivity is quantified based on the percentage of tumor cells exhibiting weak (1+), moderate (2+), or strong (3+) intensity of cytoplasmic, membranous and golgi-like staining. The results are reported as H score, obtained by the formula: 3 x percentage of strongly staining cells + 2 x percentage of moderately staining cells + percentage of weakly staining cells, giving a range of 0 to 300. A “+” sign reflects the presence of membranous staining. Patients were required to have an H-score ≥50 to be eligible for trial participation.

### Cytokine assay

CSF samples collected from the implanted Rickham catheter were analyzed using a cytokine bead array at the Clinical Immunobiology Correlative Studies Laboratory (CICSL) at City of Hope. Cytokine concentrations were quantified using the Human Cytokine 30-Plex Panel (Invitrogen) and a FLEXMAP 3D system (Luminex).

### Correlative flow cytometry of CSF and peripheral blood

Cells were washed and immunophenotyped by flow cytometry using fluorochrome-conjugated antibodies against CD3, CD8, and CD19. Data were acquired on a MACSQuant Analyzer 10 (Miltenyi Biotec) and analyzed using FCS Express software (7.28.0035) and GraphPad Prism. The gating strategy is depicted in Fig. S20.

### Human anti-CAR antibody (HACA) assay

Mock-transduced or IL13BBz-CAR–expressing Jurkat T cells were seeded at 1×10⁵ cells per well in 96-well plates. Cells were incubated with Fc receptor blocking reagent (1:20; BD Pharmingen) for 10 min, followed by incubation with participant CSF (1:3 dilution) or serum (1:10 dilution) for 1 h. A biosimilar anti-CD19 antibody (4 µg/ml; BioVision) was included in parallel wells to generate a standard curve. Cells were then stained with anti–human IgG1 Fc–Alexa Fluor 488 (20 µg/ml; Invitrogen) for 20 min, followed by staining with anti-CD3–Viogreen and anti-CD4–PerCP antibodies to enable gating. IgG1 binding was quantified using a MACSQuant Analyzer 10, with data analysis performed using FCS Express (7.28.0035) and GraphPad Prism (v10).

### Generation of TCR-reporter Jurkat cells

Thirty-nine clonally expanded TCRs from three non-lymphodepleted participants were chosen for sequencing.TCRs were cloned in plasmids and expressed in TCR-KO Jurkat T cells using published protocols^55^. Briefly, full-length TCRA and TCRB chains with murinized constant domains were synthesized in the TCRB/TCRA orientation, separated by Furin SGSG P2A linker (Integrated DNA Technologies). Gene fragments were cloned into a lentiviral vector under the control of the pEF1a promoter using Gibson assembly (New England Biolabs Inc). TCR knockout Jurkat cells expressing human CD8 and fluorescent reporters driven by transcription factor response elements and a minimal promoter (NF-kB-eCFP; NFAT-eGFP and AP-1-mCherry) were obtained from Dr. Steinberger^56,57^. Cells were cultured in RPMI supplemented with 10% FBS and 1% penicillin-streptomycin. Lentiviral particles were generated by transient transfection of the lentiviral packaging Lenti-X 293T cells (Takahara) with the TCR encoding plasmids and packaging plasmids (VSVg and PSPAX2) using Transit LT-1 (Mirus). Lentiviral supernatant was harvested 2-3 days after transfection, for 2 subsequent days, and used to transduce Jurkat cells, as described in ^55^. Lentiviral transductions were performed by spinoculation of the virus at 2,000 rpm, 37°C for 2 hours, and cells were cultured on viral supernatant for 2 days. Transduction efficiency was determined by flow cytometric analysis using the anti-mTCRB antibody^55^. When transduction of Jurkat cells was <90%, TCR-expressing cells were positively selected using anti-CD3 microbeads (Miltenyi Biotechnology).

### Functional screening of clonally expanded patient TCRs

Antigen-presenting cells (APCs) were cultured and peptide-loaded by incubating APCs with peptides at a final concentration of 1 µg/mL in serum-free medium in round-bottom 96-well plates. After 1 hour, triple parameter reporter (TPR) Jurkat cells expressing patient-derived TCRs were added. TPR Jurkat cells expressing TCR50, specific for the EBV LMP2 epitope (CLGGLLTMV), were used as a positive control, while TCR-knockout TPR Jurkat cells served as a negative control. Additional controls included TPR Jurkat cells cultured in medium alone or stimulated with αCD3/αCD28-coated beads or PMA– ionomycin. Following 20–24 hours of co-culture, cells were harvested for flow cytometric analysis. Cells were washed with PBS and stained with Zombie NIR™ Fixable Viability Dye for 15 min at room temperature in the dark. For surface antigen staining, cells were incubated with human Fc blocking reagent in for 10 min at room temperature, followed by staining with surface antibodies for 1 h at 4 °C. Cells were then washed with staining buffer (2% FCS and 2 mM EDTA in PBS) and fixed using IC Fixation Buffer according to the manufacturer’s instructions. Stained cells were stored at 4 °C overnight and both acquired and analyzed the following day on a NovoCyte Quanteon cytometer (instrument configuration 4025) using NovoExpress software (V1.6.2.). Data visualization was performed using GraphPad Prism (v11). A schematic of the gating strategy is provided in Fig. S20.

### Single cell RNA and whole exome library preparation and sequencing

Single-cell library preparation was carried out on 33 clinical patient derived samples from 5 patients according to 10X Genomics Chromium Next GEM Single Cell 5’ Reagent Kits v2 (Dual Index) CG000331 Rev A. 13 Peripheral blood mononuclear cells (PBMC), 5 CAR-T products, 5 tumors, and 10 CSF samples were placed into 6 batches yielding GEX, TCR V(D)J, feature barcode, and CAR transcript enrichment libraries (**Table S5**). Sequencing was achieved on the NovaSeq6000. An additional 11 batches were run on 34 patient derived samples from 6 patients using an updated 10x protocol *Chromium GEM-X Single Cell 5’ Reagent Kit V3 CG000733 RevA*. Batches 1-6 contained 2-3 clinically relevant timepoint CSF patient samples. Batches 7-11 contained 2-3 multiplexed PBMC samples (**Table S5**). GEX, TCR, and CAR libraries were produced for batches 1-11, with 4 nested primers (RT spike in) being used for every CAR library. GEX, TCR and CAR libraries were sequenced on the NovaSeqXPlus platform per 10x sequencing recommendations.

Batches 37, 38, 39, and 40 used Biolegend Total Seq C Human hashtag antibodies to barcode product and PBMC samples and allow for later patient pooling and deconvolution^58^. PBMC, CAR-T product, and tumor samples were sorted for viability using -AM live stain and sorted into a collection tube for further processing. All CSF and tumor samples, and 1 CAR-T product, were loaded directly to individual GEM reactions. CSF samples did not undergo Calcein-AM staining or preprocessing and were loaded to GEM directly following cryopreservation thaw. CAR transcript enrichment libraries were performed on batch 37 with a series of 3 custom nested primers starting at cDNA amplification, then subsequent batches used a series of 4 nested CAR primers included at reverse transcription to further enrich lowly transcribed targets. In addition, batches 37 and 38 had 3 non-hashed CSF samples run in a separate Gel Bead-in-Emulsion (GEM) reaction. Batch 42 included 4 non-hashed CSF samples, 3 duplicate samples from batch 37 and one new patient CSF sample. Duplicate libraries were generated in order to saturate CAR transcript diversity. Batch 39 contained 4 hashed samples and 2 non-hashed tumor samples. Batch 40 contained 3 hashed samples and 1 non-hashed tumor sample and 1 non-hashed product sample. Batch 41 contained no hashing and 2 non-hashed tumor samples. Hashed live single cell patient samples were pooled at 10,000 cells per sample and loaded to GEM. Multiplexed GEM reactions were pooled and loaded at 30-50k cells per GEM to maximize yield and restrict unidentifiable doublet counts using Satija Lab Multiplexing Cost Calculator^59^. Cell count validation using Illumina Iseq100 provided a recommended sequencing depth for the relevant library type and pooled accordingly to NovaSeq6000.

Alternative to cell hashing, demultiplexing was performed on batches 1-11 using patient whole exome profiles. Whole exome sequencing was performed with twist exome 2.0. Each patient sample was sequenced at 100x coverage on The NovaSeqXPlus platform. Whole exome data was processed, and variants were called per sample using TGen’s Tempe v1.2.2 pipeline, part of the Jetstream workflow, using the default parameters (https://github.com/tgen/tempe).Gene expression libraries were run with CellRanger v5.0.1 for batches 37-42 using the count function and default settings ^60^. GEM-X chemistry is not supported in CellRanger v5.0.1 so batches 1-11 were run with CellRanger v8.0.1 using the same settings. GRCh38 Ensembl 98 with the CAR construct added was used as the reference. For the samples that were multiplexed (**Table S5**), we used the same approach as above but added the -- include-introns flag and provided the antibody panel (Total Seq C).

### Place holder – clinical counts for PBMC and CSF

Using these clinical count data, we calculated lymphocyte count in the CSF. Lymphocyte counts were calculated by multiplying total counts by the lymphocyte percentage and rounding to the nearest whole number. We plotted both lymphocyte counts for PBMC and CSF using ggplot2.

### scRNA-seq data analysis

Single cell gene expression matrices were processed in R v4.4.0 ^61^ using Seurat v5.1.0^62^. Sample-type specific filtering was performed for the gene expression matrices using the following filtering thresholds 1) CSF: pt_mt = 10, nFeature = 650, nCount = 1200, 2) PBMCs: pt_mt = 10, nFeature = 650, nCount = 1200, 3) product: pt_mt = 10, nFeature = 1300, nCount = 2500, and 4) tumors: pt_mt = 10, nFeature = 1500, nCount = 2300 (**Fig. S19**). For samples that were multiplexed (**Table S5**), we demultiplexed either using HTODemux in Seurat (for samples without exome data) or Souporcell v3 (for samples with exome data)^63^. The exome VCFs were first merged by batch using bcftools v1.10.2^64^ and then filtered using VCFtools v1.10.2^65^. to remove sites with a minor allele frequency less than 0.05 (--maf 0.05) and sites with any missing data (--max-missing 1.0) and kept biallelic sites only (--min-alleles 2 –max-alleles 2). We additionally only included genotypes that had high coverage in the scRNA data. Sites had to have at least 3000 reads in the scRNA data to be included in the analysis. We then ran Souporcell with the standard parameters. Two samples (UPN705, batch 3 and UPN689, batch 8) failed souporcell due to low Person correlation. Those samples were removed and Souporcell was run again (**Table S14**). After demultiplexing, Seurat objects were converted to 10x sparse matrices using DropletUtils write 10XCounts^66^. We next estimated and corrected the gene expression matrices for ambient RNA using SoupX v1.6.2^66,67^. Contamination fraction was automatically estimated for each sample using the autoEstCont function and gene expression matrices were corrected using the adjustCounts function in SoupX. Cell cycle scores were then assigned using the CellCycleScoring function in Seurat and regressed out during normalization. We ran DoubletFinder v2.0.6^68^ on each batch to identify doublets using a doublet rate of 15%. We loaded cells with a desired doublet rate of 15%. Doublets were removed prior to integration. Normalization was performed using SCTransform v2^69,70^. Mitochondrial and ribosomal genes were removed prior to integration and downstream analyses.

We used the rPCA method for integration followed by PCA dimensionality reduction on the top 2500 most variable genes and visualization using Uniform Manifold Approximation and Projection (UMAP). The number of PCs used in the UMAP visualizations were determined by selecting the PC where change of percent of variation is less than 0.1%. Nearest-neighbor graphs were constructed using the PCA reduction and cells were clustered to various resolutions using the Louvain algorithm. Data were integrated by sample - UPN/Cycle/day/batch combination for CSF and PBMC, UPN for product, and UPN/batch for tumor. CSF, PBMCs, product, and tumors were integrated separately, and CSF and PBMC samples were integrated together.

Cell type annotations were performed by visualizing gene expression of known marker genes and top markers for each cluster (**Table S15, Fig. S4-S8, S18**). Sub-clustering was performed where needed using FindSubCluster function in Seurat. Top markers were identified using Presto’s v1.0.0 Wilcoxon rank sum test to identify top markers for each cluster ^71^. Myeloid and lymphoid cells were identified then re-integrated separately before proceeding to cell type annotation.

T cell states were further annotated for CSF, PBMC, and product. We first subset T cell clusters from PBMC and CSF lymphoid objects and performed integration, dimensionality reduction, clustering, and visualization as described above. Clusters with expression of non-T cell genes (for example, *LYZ*) were removed, data were re-clustered, and the UMAP was re-generated. Similar to the cell type annotations, T cell states were identified by visualizing gene expression of known marker genes and top markers for each cluster (**Table S15**, **Figures S4-S8, S18**).^72^ We additionally annotated CAR positivity in all T cells. CAR^+^ T cells were identified as an annotated T cell having three or more reads supporting the CAR construct (IL13OP) while all other T cells were categorized as CAR negative.

Cell type proportion differences were tested and visualized using scProportionTest v0.0.0.900 ^73^, dittoseq v.1.8.1^74^, and ggplot2. scProportionTest facilitates permutation tests to obtain p-values for each cluster and bootstrapping to obtain confidence intervals to compare the differences in proportions of cells between scRNA-seq samples. Patients were grouped whether they received lymphodepletion prior to treatment or not. We also stratified our cell type proportion analysis by early and late timepoints (**Fig. S10**). Early time point was defined as the first cycle we had scRNAseq for that was less than cycle 5. We did not include an early cycle for UPN705 because the earliest cycle we had for that patient was cycle 5. Late time point was defined as the last cycle we had scRNAseq for.

To assess whether expression in PBMC is a good proxy for expression in CSF, we tested whether sample level expression in PBMC is associated with sample level expression in CSF (**Fig. S12**). Linear models were run in R, where gene expression in CSF samples was the outcome variable and gene expression in PBMC samples was the predictor variable (lm(CSF ∼ PBMCs)). For each sample and cell type we pseudobulked by calculating the mean counts for all of the cells for each UPN, cycle, and compartment (CSF or PBMC) combination. Any sample with less than 10 cells for a cell type were not included in the analysis and any cell type with less than three PBMC-CSF matched samples were excluded. We additionally removed genes where at least half of the samples had no expression in each sample type. FDR correction was performed on each cell type.

### scTCRseq data analysis

scRepertoire v2.0.7^75^ was used on the filtered contig annotation files to identify clonotypes. We required both alpha and beta chains to be present and used the strict calling method (VDJC gene and CDR3 nucleotide present) when calling clonotypes. TCR frequency (total number of observed cells with the clonotype) were calculated for each UPN, cycle, and sample type combination. Clonotypic information generated from scRepertoire was added to the single cell Seurat objects using the combineExpression function. TCR frequencies were normalized by the total number of T cells for each UPN, cycle and sample type combination. Expanded TCRs were defined as having a TCR count greater than one and having a normalized TCR frequency in the top 20% of normalized TCR frequencies across all samples. We further categorized TCRs as “expanded” with a normalized TCR frequency between the top 20% to 5% of the distribution, “more expanded” with a normalized TCR frequency between the top 5% to 1% of the distribution, and “most expanded” with a normalized TCR frequency at the top 1% of the distribution.

### CSF cell-free DNA sequencing and variant allele fraction analysis

Variant allele quantification of patient CSF was accomplished via targeted sequencing of known tumor mutations. For each patient, tumor associated mutations were identified from clinical diagnostics performed during care. Multiplex PCR primer sets were then designed for all known clonal mutations (1-3 primer sets per patient) and validated for specificity via gel electrophoresis. Cell free DNA was extracted from 0.5ml - 1ml of CSF using the Qiagen QIAamp MinElute ccfDNA kit (QIAGEN #52284). To maximize recovery of cftDNA and increase sensitivity, extracted DNA was not quantified and immediately vacuum concentrated at 35°C to <20ul (Savant SpeedVac DNA130). The entirety of each DNA extract was then subjected to 40 cycles of multiplex PCR amplification (primers and cycling parameters to be supplied after publication) using NEB Q5 HotStart 2x master mix (NEB #M0494) with primers at 0.5uM final concentration. PCR product was then cleaned, barcoded, and sequenced via Oxford Nanopore MinION according to the manufacturer’s instructions (Oxford Nanopore #SQK-NDB114.24) and basecalled using dorado v1.1.1 leveraging the “super” basecalling model (sup@5.2.0). Reads were aligned using minimap2 v2.24 (-x map-ont -n 2 -m 20) and allele fractions were computed by inspecting allele counts via Integrated Genome Viewer (IGV v21.0.4; VAF%=mut/(mut+wt)*100).

### Statistical analysis

Median overall survival from first infusion and median overall survival from diagnosis along with 95% confidence limits are reported for patients receiving the four required CAR T cell infusions by diagnosis and treatment. Counts are presented for adverse events by treatment and diagnosis. Counts and percentages are presented for disease response (RAPNO) by diagnosis and treatment. This work was conducted using R v.4.5.2 with Rstudio v.2026.01.1.

GraphPad Prism (GraphPad Software) was used to generate bar plots and graphs.

## Data Availability

Scripts for processing sequencing data will be made publicly available on GitHub at https://github.com/Banovich-Lab/19130_Pediatric_CART. Aggregated patient data, once deidentified, will be made available upon request. TCRseq and scRNAseq data from patient samples reported here will be deposited in GEO under the accession number GSE261235. Sequencing data (FASTQ files) for the CSF VAF studies will be deposited in an appropriate repository upon manuscript acceptance/final manuscript preparation. An accession/link will be provided in the final submission. Until deposition, data will be made available from the corresponding author upon reasonable request, subject to any applicable institutional and patient-privacy restrictions.

## Acknowledgments

First and foremost, we thank our patients and their families, who were invaluable and incredibly generous partners to our team. It was a tremendous honor to be part of their journey. We are also very grateful to Michelle Monje, Sohel Talib, Lisa McGinley, Abla Creasey, and other members of the CIRM Clinical Advisory Panel for their support of this trial. We thank other members of the Leo Wang Lab and the T Cell Therapeutics Research Laboratories for their discussion and assistance. Research reported in this publication included work performed in the GMP Manufacturing, Analytical Cytometry, Analytical Pharmacology, and Research Pathology Services Shared Resources supported by the National Cancer Institute (NCI) of the National Institutes of Health (NIH) under grant number P30CA033572. Work done by the Children’s Hospital of Los Angeles Center for Pathology Research Services and the USC Norris Cancer Center Translational Pathology Core in obtaining and preparing surgical tumor samples was supported by NCI grant P30CA014089. Other federal support included the National Institute of Neurological Disorders and Stroke (R01CA155769, B.B., C.E.B.) and the Food and Drug Administration (R01FD005129, C.E.B.). Work was also supported by grants from the California Institute for Regenerative Medicine (CLIN2-12153 and INFR4-13587, L.D.W.), the V Foundation (T2022-004, L.D.W.), Gateway for Cancer Research (G-15-600, C.E.B.), the Marcus Foundation (L.D.W; C.E.B.), the Turtle Pond Foundation (L.D.W.), the Panda Cares Foundation (L.D.W.), the ChadTough Defeat DIPG Foundation (C.K.; D.D.N; L.D.W.; including the Mark Massey Legacy Foundation and Elle’s Angels Foundation to L.D.W.), and the Pediatric Cancer Research Foundation (L.D.W.). C.J.W. and K.M.C. are members of the Parker Institute for Cancer Immunotherapy. C.J.W is supported in part by U24CA224316 (NIH/NCI). C.J.W. is also the Lavine Family Chair for Preventative Cancer Therapies at DFCI. The content is solely the responsibility of the authors and does not necessarily represent the official views of funding agencies, including the National Institutes of Health.

## Author Contributions

L.D.W. is the principal investigator of the trial. T.B.D. and S.S. are the site investigators. C.E.B. and S.J.F. are the Investigational New Drug (IND) holders. L.D.W., C.E.B., J.W., and B.B. conceived the project. J.W. and C.E.B. oversaw regulatory affairs and manufacturing. L.D.W., C.E.B., M.S.B., J.W., M.N., M.H., A.M.D., J.L.K., T.B.D., and B.B. planned, designed, and wrote the clinical trial. L.D.W., M.M., S.S., J.G., B.B., L.F., J.S., T.B.D., S.A.R., C.K., S.A.U., J.A.R., and J.L.K. were responsible for patient care. B.T. and J.R. reviewed radiographic studies and defined responses. G.S., S.M.S., J.C.H., M.A., L.P., A.T.O., R.V.M., H.M.N., G.O., K.L., Y.V., S.E.L., and M.W. participated in processing of patient samples and/or correlative studies. T.S. and S.B. performed statistical analysis. L.P., A.T.O., R.V.M., H.M.N., M.W., K.M.C., D.G.C., and N.E.B. carried out single cell processing, sequencing, and analysis. J.C., L.A., and M.D. performed pathologic evaluation of patient samples, including immunohistochemical analysis. C.K., J.W., D.D.N., and T.A. performed ctDNA studies. L.D.W., N.E.B., and C.E.B. oversaw analysis of all correlative studies. L.D.W., A.T.O., and S.E.L. wrote the manuscript. All authors reviewed and edited the manuscript. L.D.W., N.E.B., and C.E.B. supervised all aspects of the work.

## Supplemental Figures

**Fig. S1.**
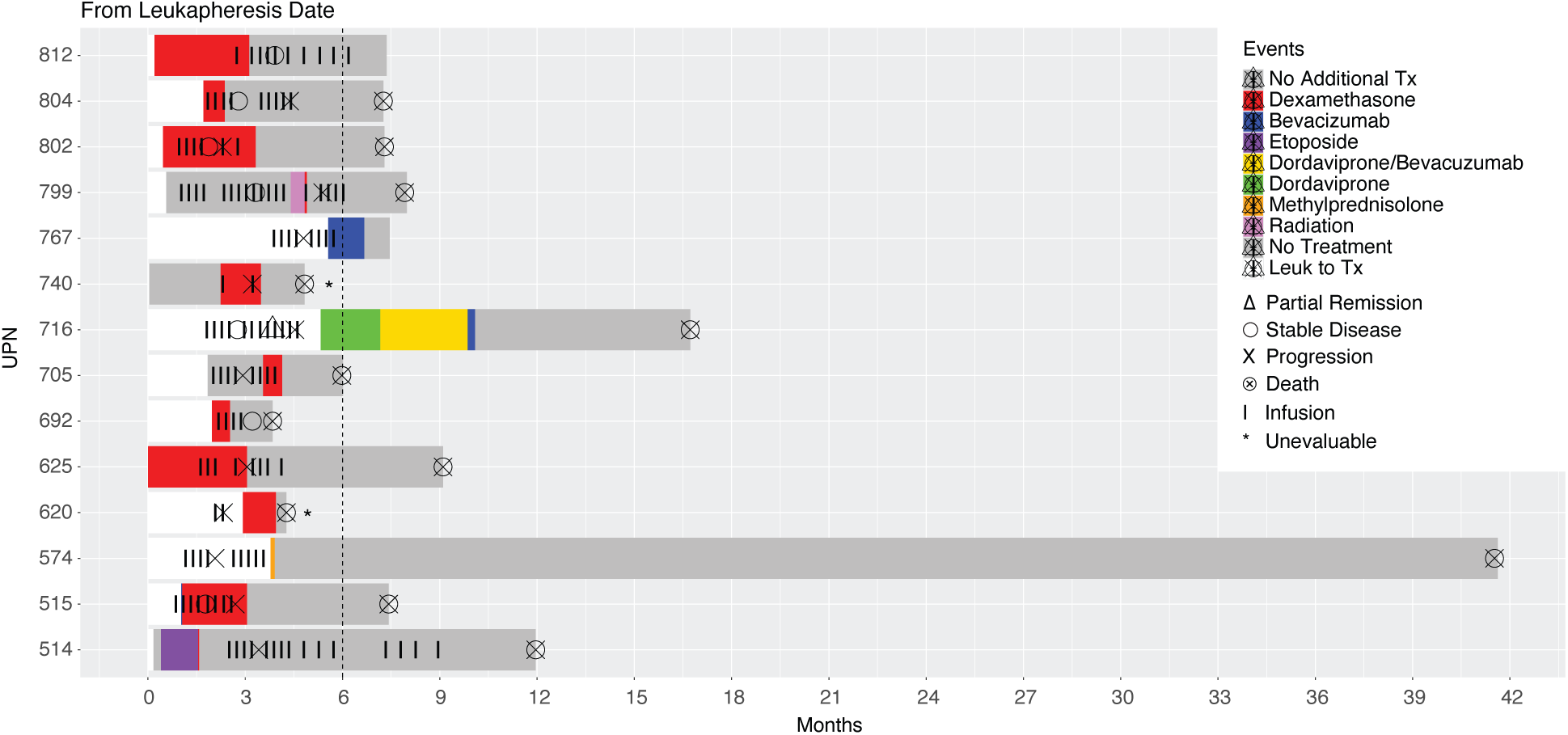
Anti-inflammatory and antitumor therapies concomitant to CAR T cell therapy. Swimmer plot timeline of concomitant drug treatments for patients relative to leukapheresis and CAR T cell infusion. Patients who did not use concomitant therapies are not shown. Colored bars denote periods of treatment for each therapy. Vertical black bars denote CAR infusions. Wireframe symbols indicate disease response status.

**Fig. S2.**
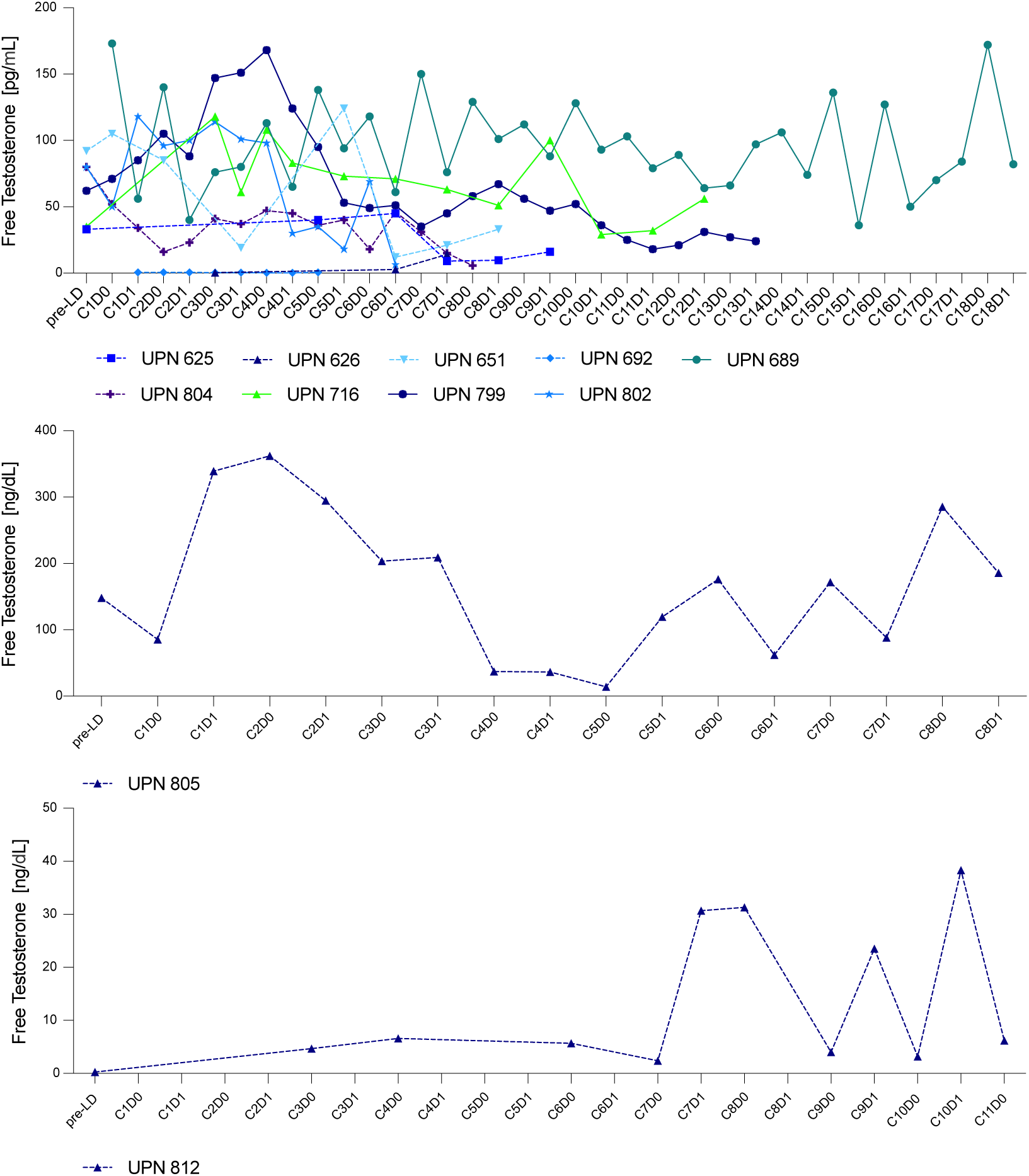
Testosterone over the course of therapy in male patients. After September 2022, free testosterone levels were measured in the clinical laboratory and reported at each visit for male patients. Patients ranged in age from 14 to 22. Four patients were noted to have pre-treatment testosterone levels <50ng/dL. All other patients experienced fluctuation in their testosterone levels over the course of therapy. UPN805 and UPN812 were measured in different clinical systems from the other patients, so their levels are represented individually.

**Fig. S3.**
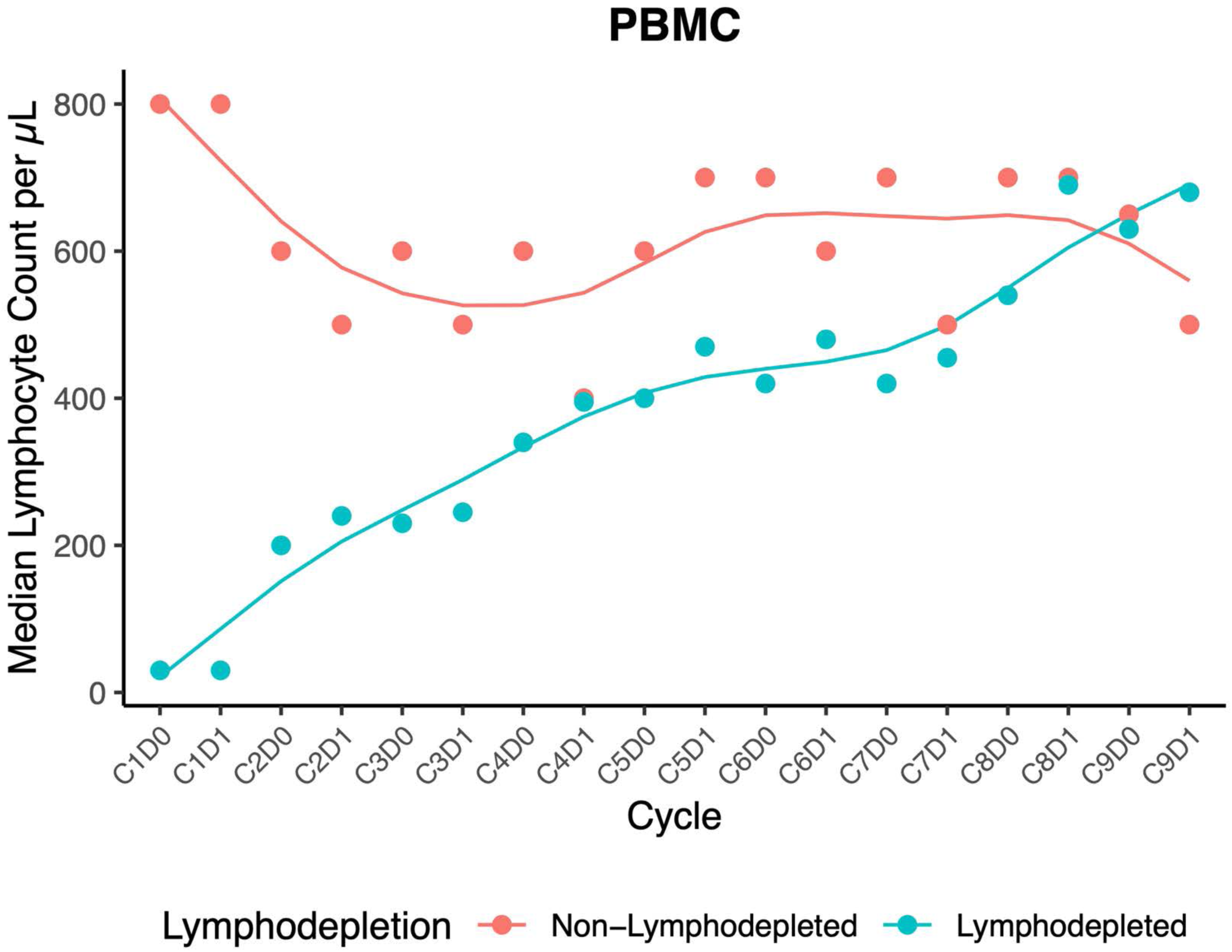
Median lymphocyte counts by patient lymphodepletion status in peripheral blood over the course of therapy. Median values (points) are shown for each cycle and day. Solid lines represent a cubic smoothing spline interpolation (smoothing parameter = 0.5) to highlight trends across treatment groups.

**Fig. S4.**
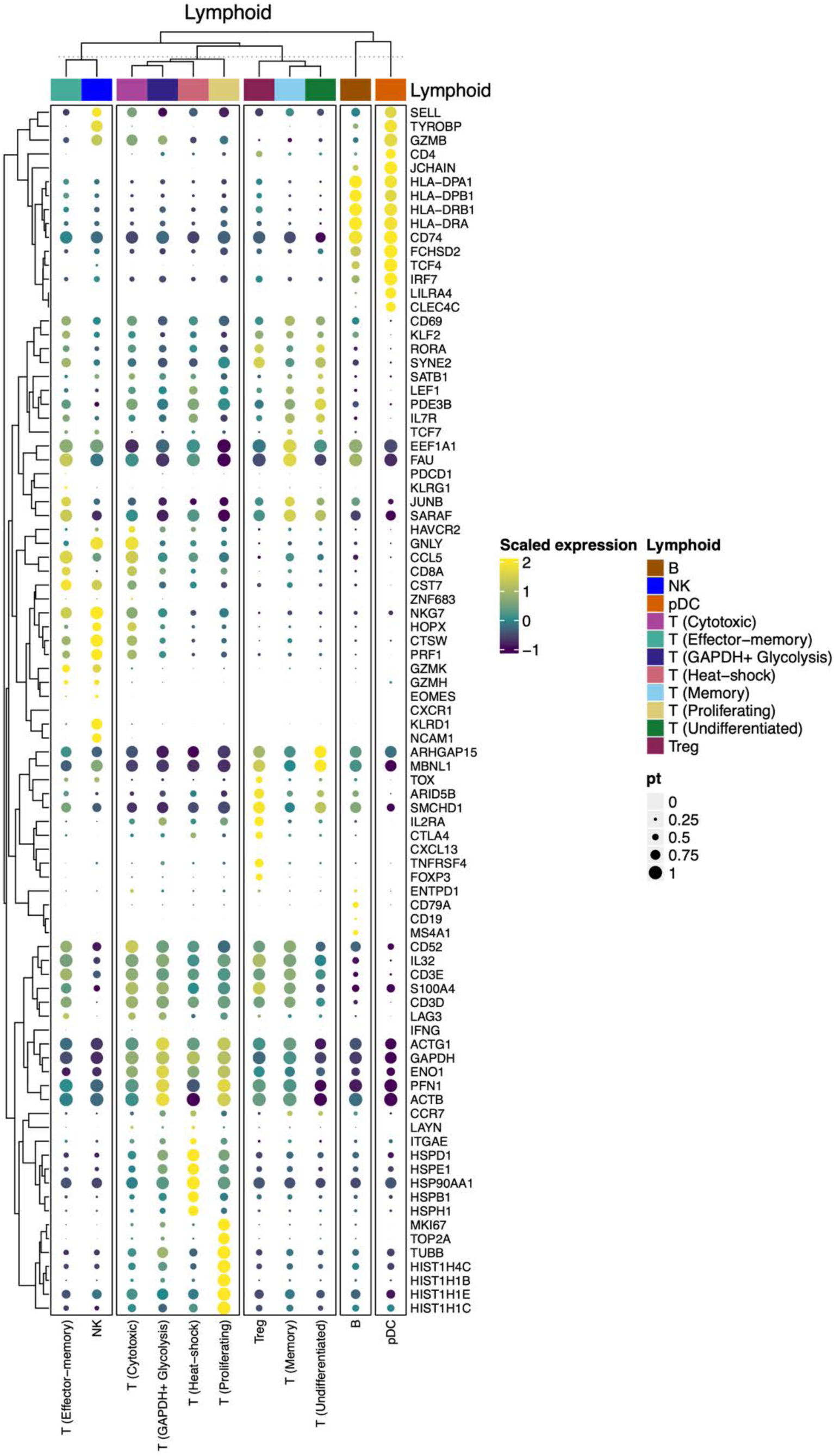
Dotplot heatmap of genes used in cell type annotations for lymphoid cells in the CSF. Top 5 markers were identified for each cell type/state along with a curated set of canonical markers.

**Fig. S5.**
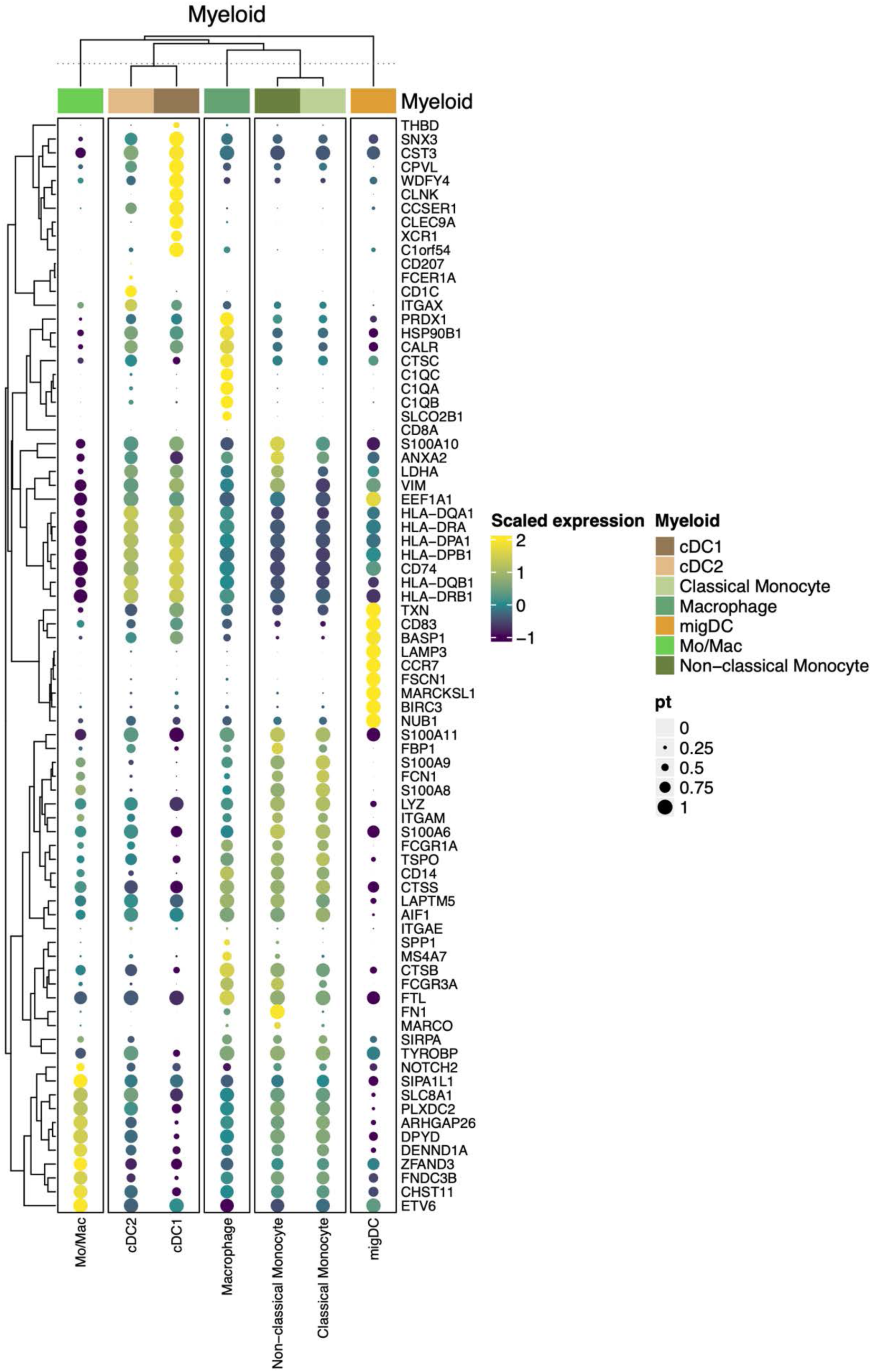
Dotplot heatmap of genes used in cell type annotations for myeloid cells in the CSF. Top 10 markers were identified for each cell type/state along with a curated set of canonical markers.

**Figure S6.**
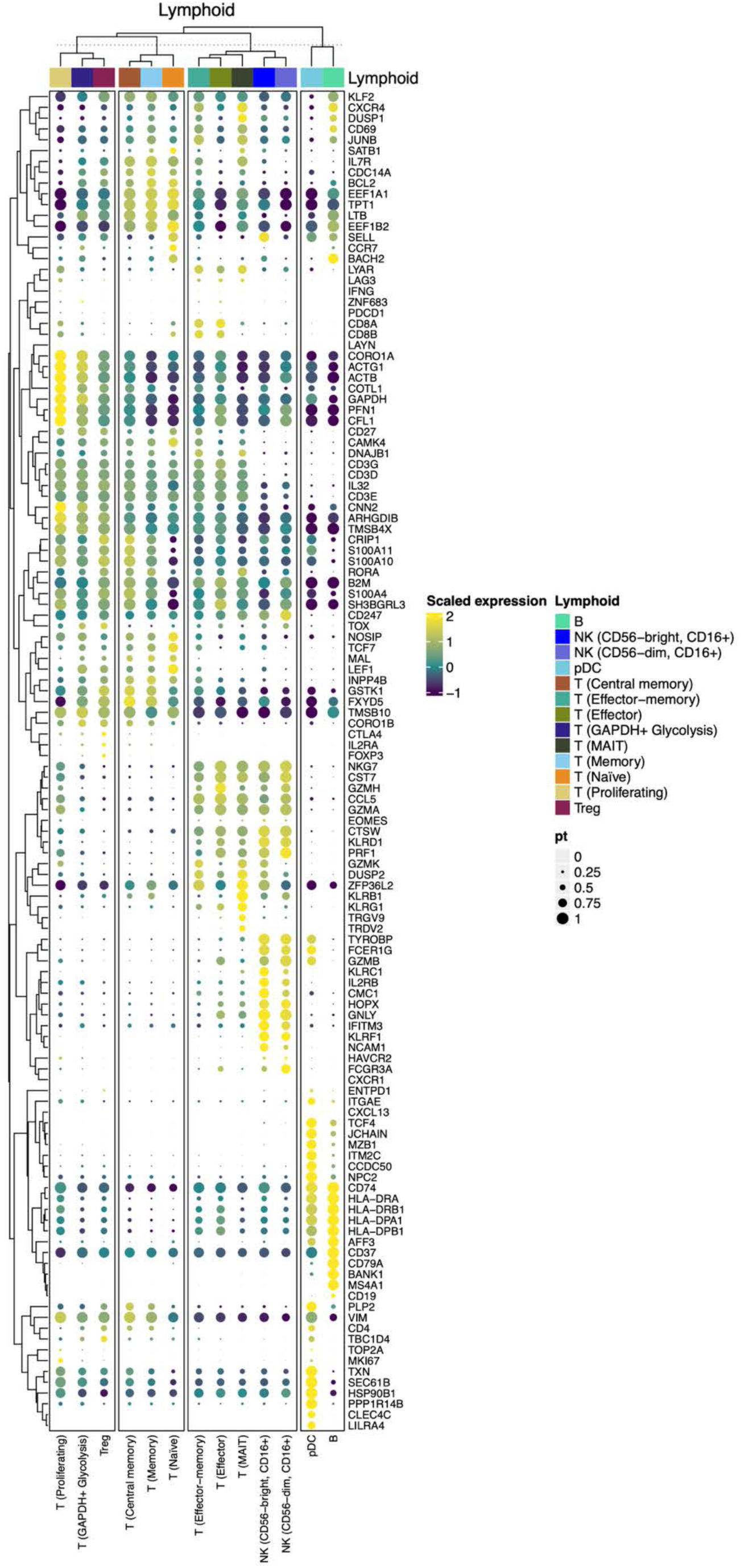
Dotplot heatmap of genes used in cell type annotations for lymphoid populations in PBMC. Top 10 markers were identified for each cell type/state along with a curated set of canonical markers.

**Fig. S7.**
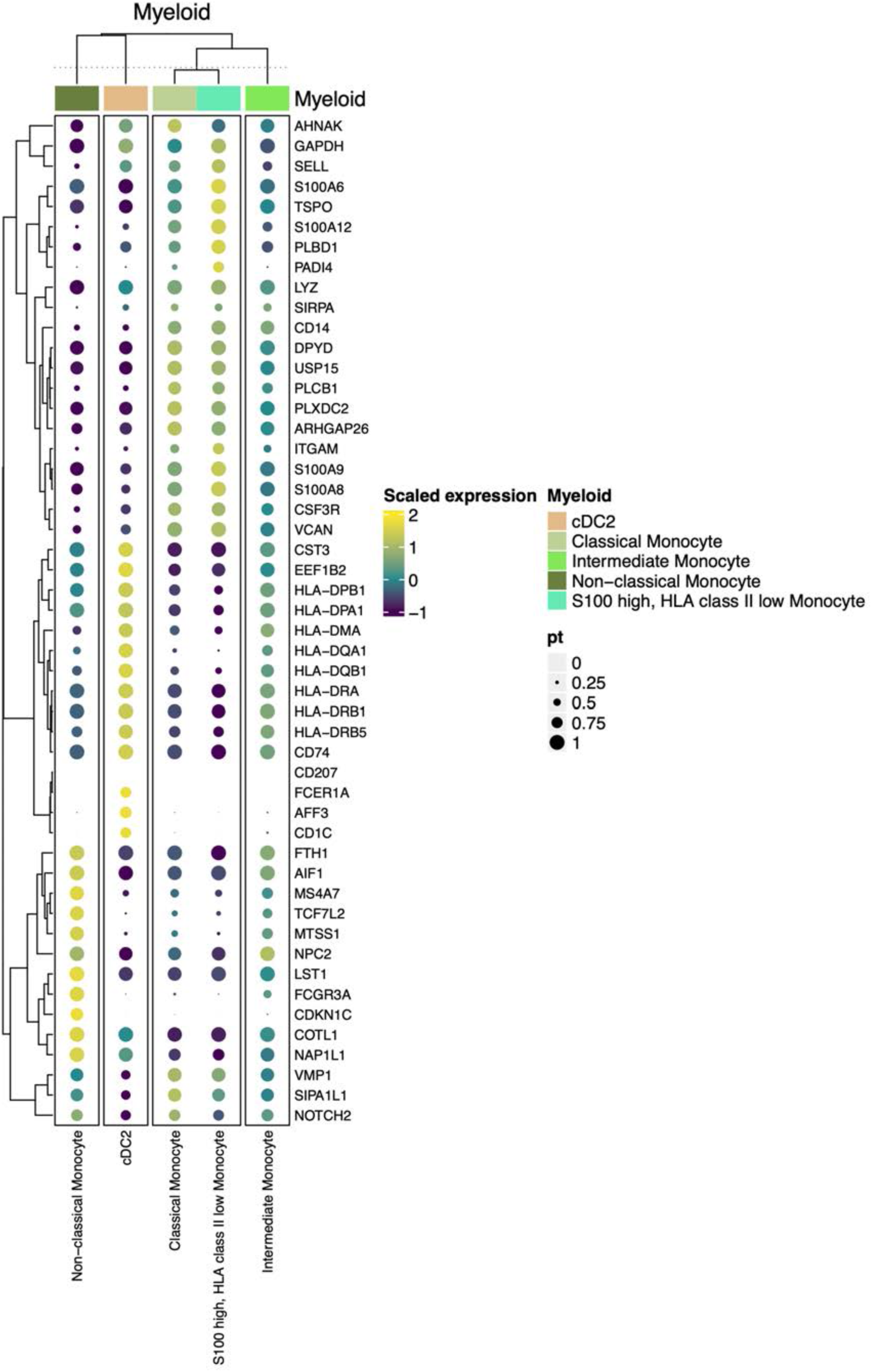
Dotplot heatmap of genes used in cell type annotations for myeloid cells in PBMC. Top 10 markers were identified for each cell type/state along with a curated set of canonical markers.

**Fig. S8.**
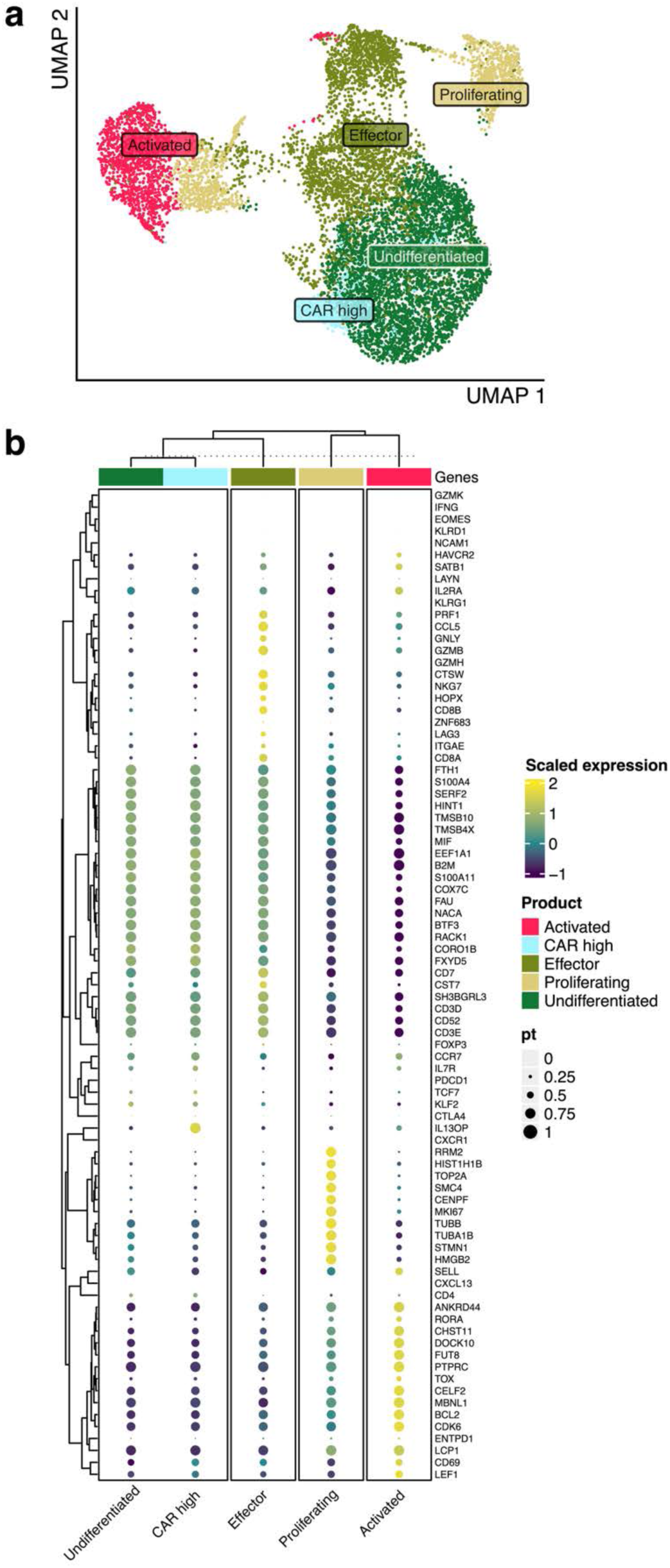
Product UMAP and dotplot heatmap of genes used in cell type annotations for T cell states in engineered product. A) UMAP of 10,885 T cells in the engineered product colored by T cell state. B) Dotplot heatmap of top 10 markers that were identified for each cell type/state along with a curated set of canonical markers.

**Fig. S9.**
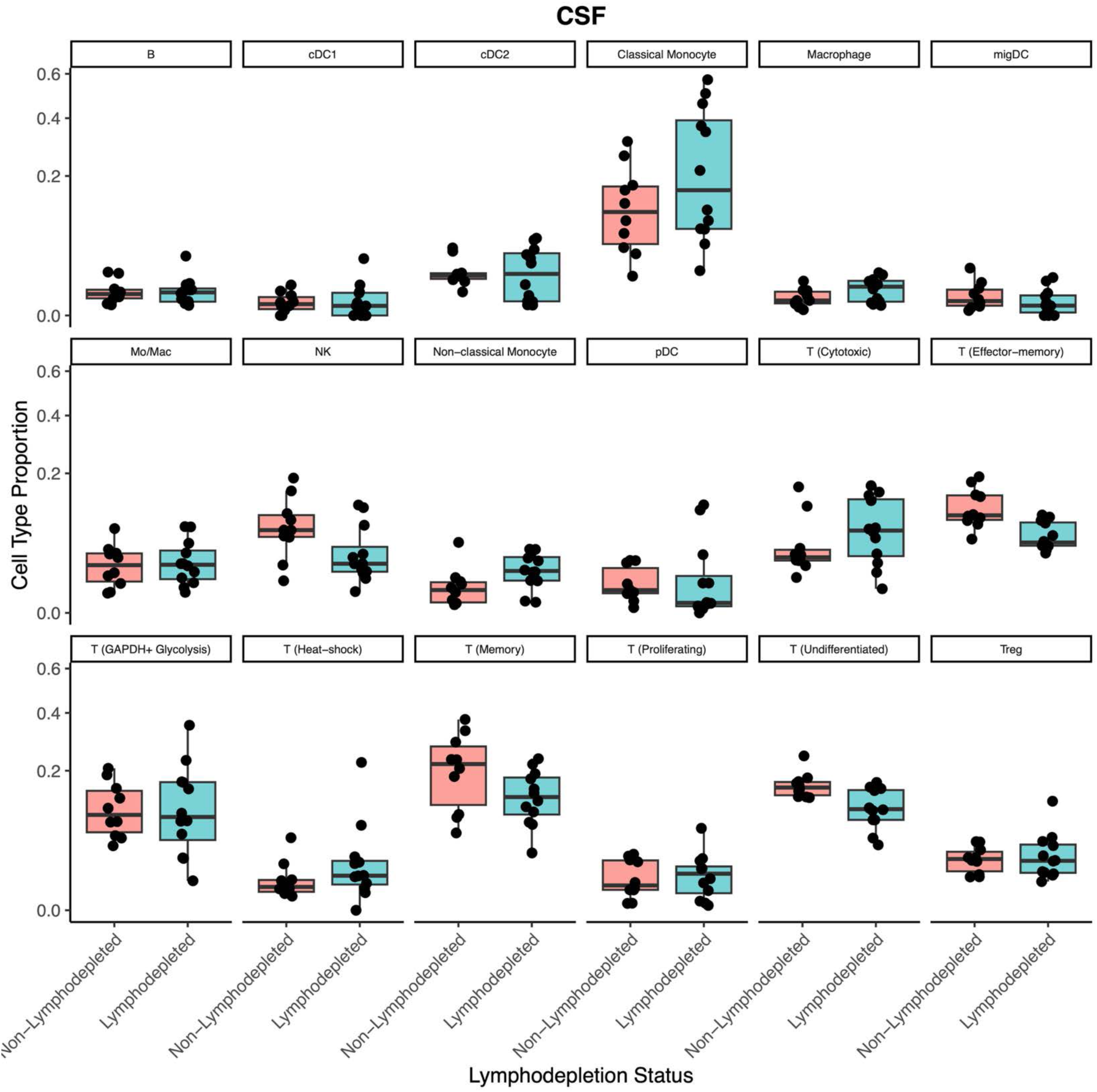
Cell type proportions in CSF. Per cell type boxplots of cell type proportions in the CSF. The y-axis was square root transformed to enable visualization of cell types with smaller proportions. Center lines represent the median; box limits represent the 25th and 75th percentiles; whiskers extend to 1.5× the interquartile range.

**Fig. S10.**
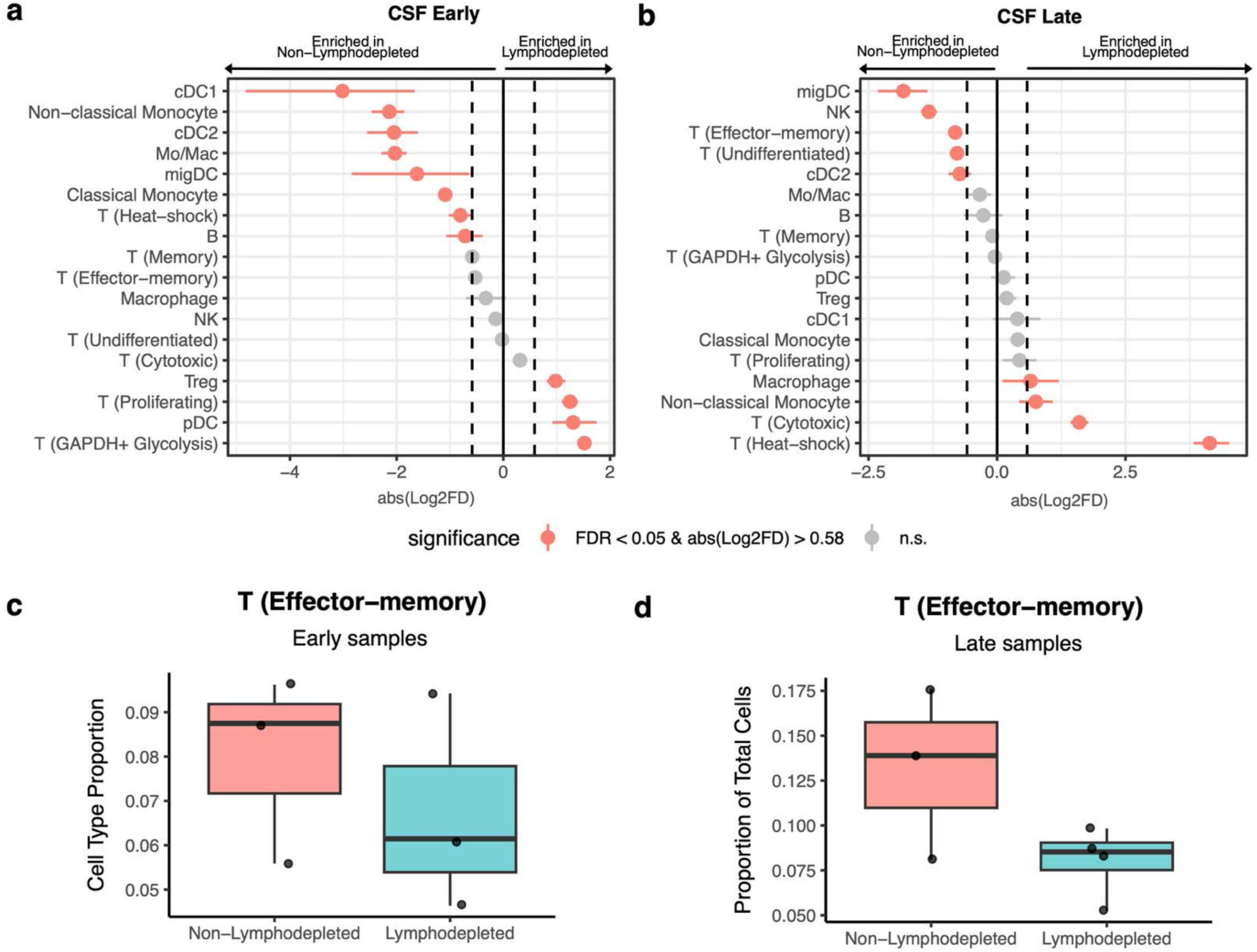
Cell type composition between lymphodepleted and non-lymphodepleted patients during early and late treatment cycles. Point range plots comparing the proportion of cell types between patients that received lymphodepletion and patients that did not during **(a)** early and **(b)** late treatment cycles in CSF. Red points in **a-b** represent cell types with a significant proportional difference between categories and the red horizontal lines represent 95% CIs. Dashed vertical lines represent absolute log2 fold difference (abs(Log2FD)) of 0.58. Significant proportional differences between categories were ones with both FDR <0.05 and abs(Log2FD) of 0.58. Solid vertical line represents abs(Log2FD) of 0. abs(Log2FD) greater than 0 represents cell types at a higher proportion in patients that received lymphodepletion compared to those who did not. Boxplots of effector memory T cell proportions between patients that received lymphodepletion and patients that did not during **c,** early treatment and **d,** late treatment cycles. Center lines in **c-d** represent the median; box limits represent the 25th and 75th percentiles; whiskers extend to 1.5× the interquartile range.

**Fig. S11.**
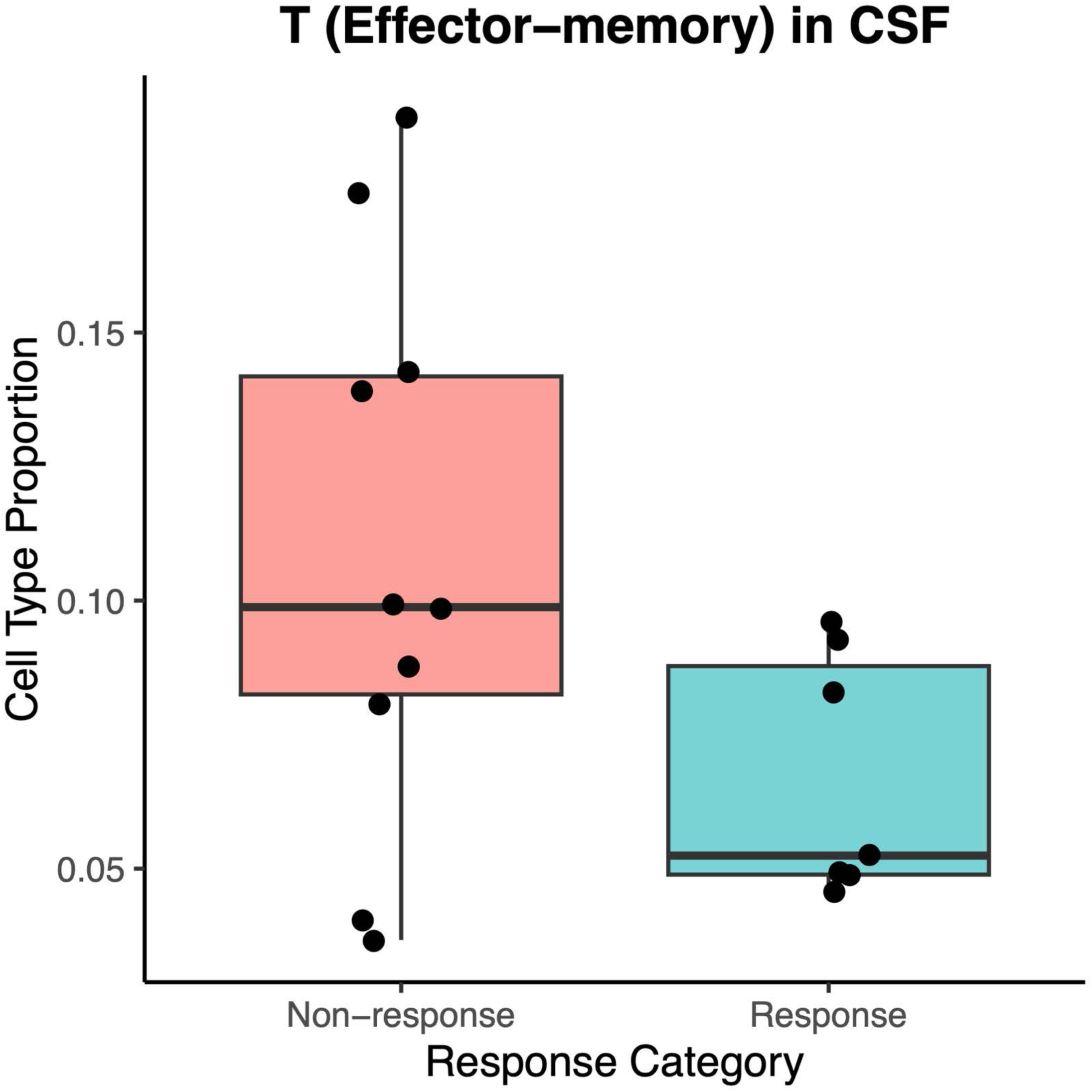
CSF effector memory T-cell abundance by radiographic response. Box plots show the proportion of effector memory T cells during periods of response and non-response. Center lines represent the median; box limits represent the 25th and 75th percentiles; whiskers extend to 1.5x the interquartile range.

**Fig. S12.**
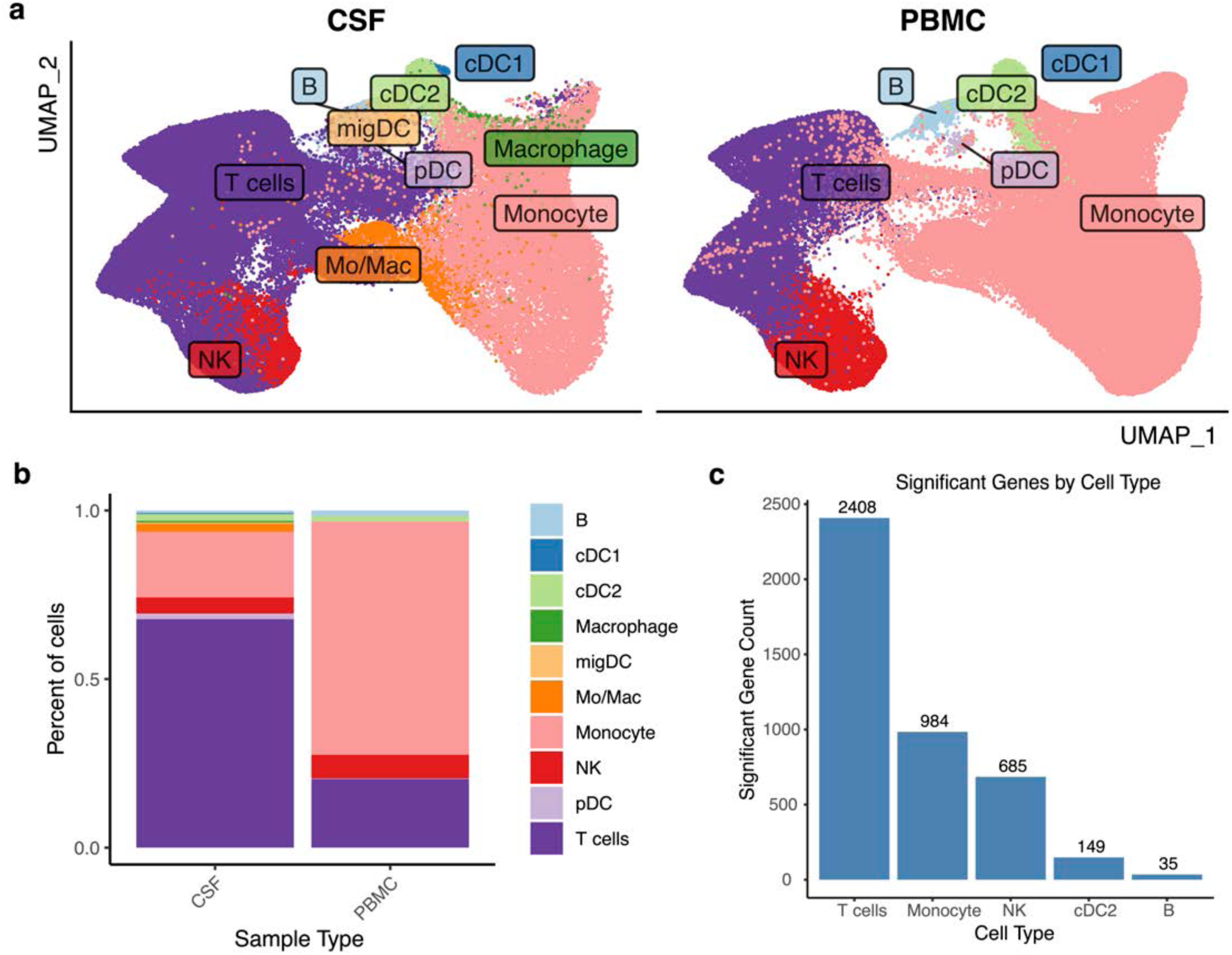
Comparison of cell type proportions and gene expression between CSF and PBMC samples. A) UMAP of CSF + PBMC integrated object split by sample type. Cell type annotations were unified between the two sample types. B) Stacked bar plot of cell type proportions for CSF and PBMC. C) Bar plot of genes with significant association of expression between CSF and PBMC split by cell type.

**Fig. S13.**
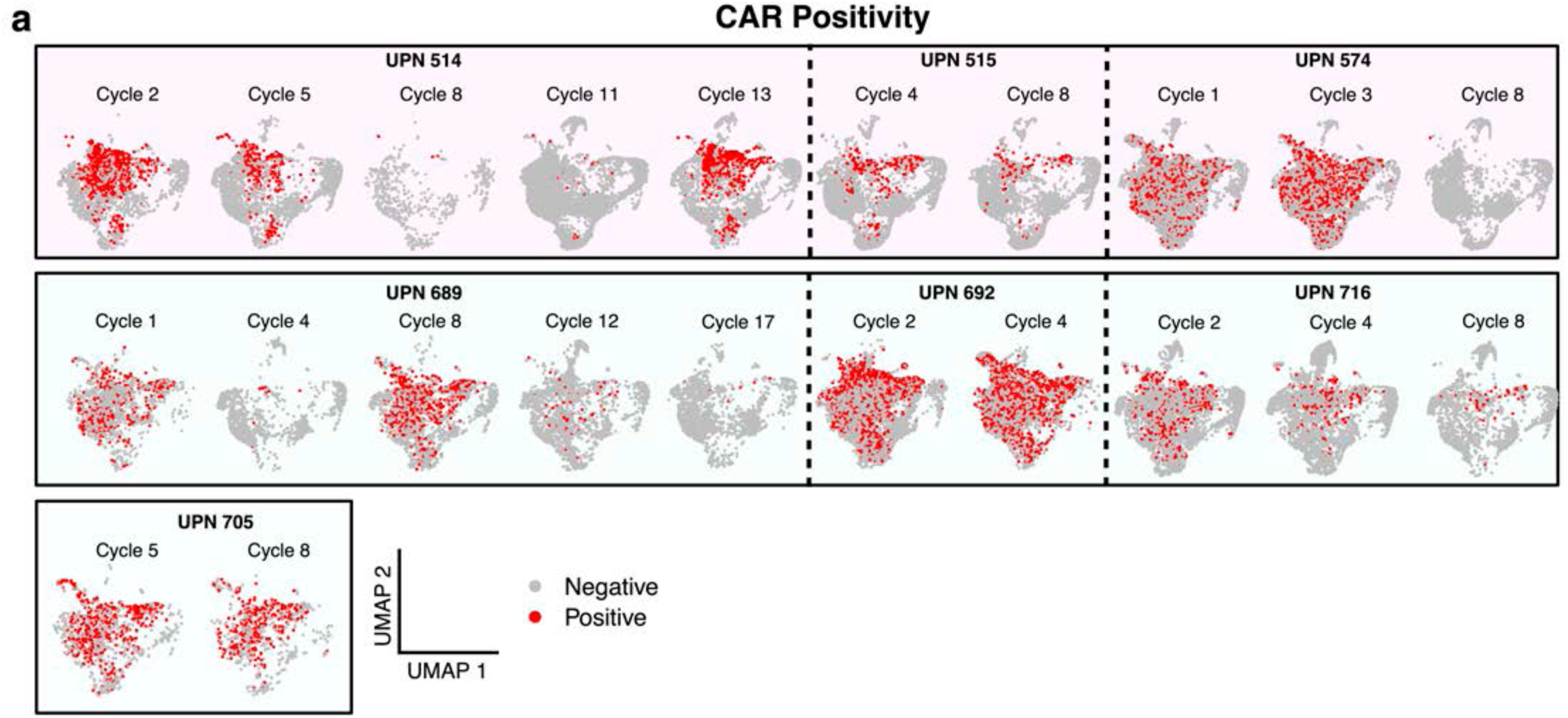
CAR T dynamics over the course of therapy. A) UMAP of CSF lymphoid cells split by patient (UPN) and cycle. Cells are colored by CAR positivity – red dots are CAR+ T cells, grey are all other lymphoid cells. B) Line plots of the proportion of CAR+ T cells over all T cells across cycles. Each color represents a patient (UPN) and the plot is split by samples that received lymphodepletion (left) and ones who did not (right).

**Fig. S14.**
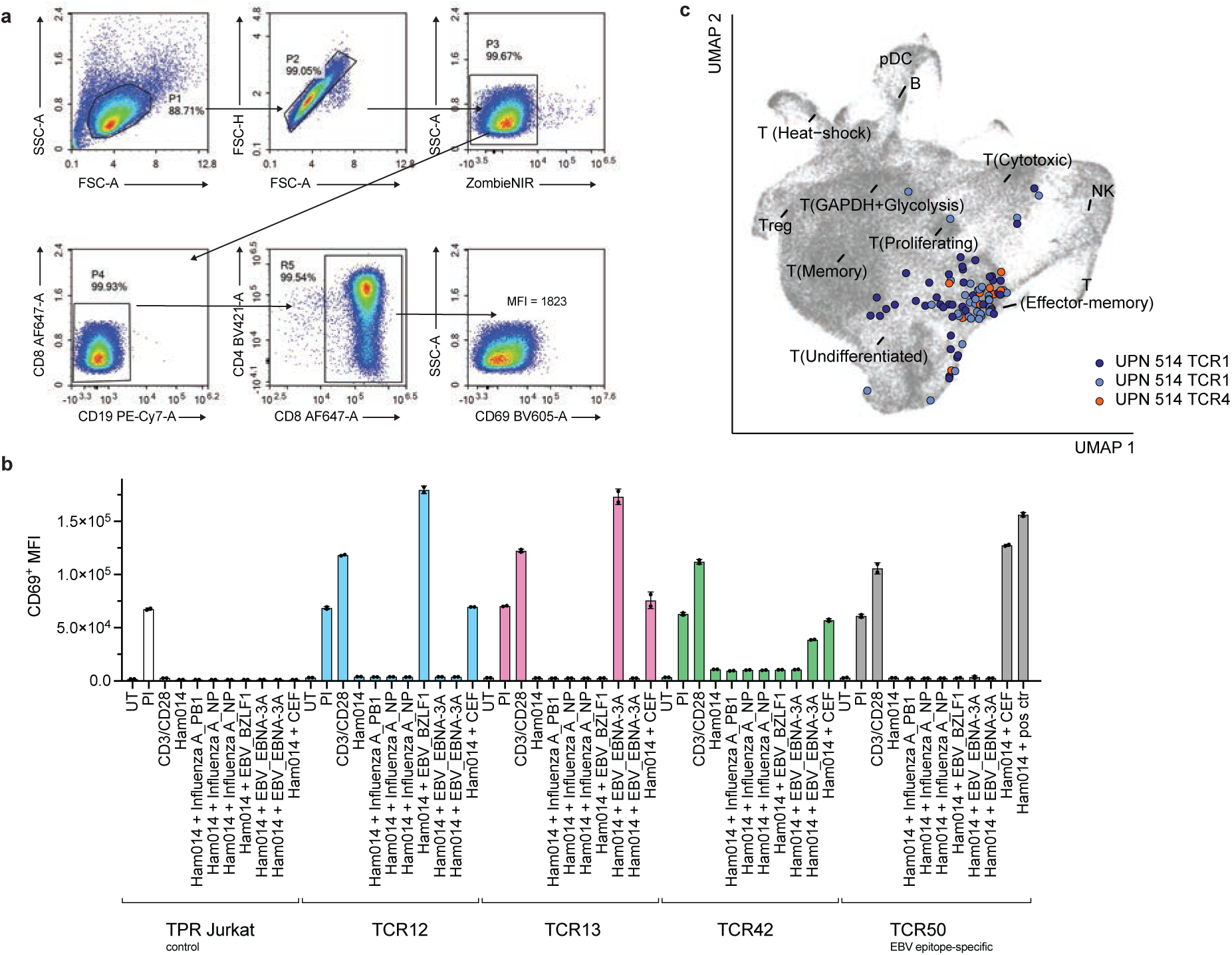
Identification of T cells recognizing EBV peptides in UPN514. **a**. Representative gating strategy for evaluating CD69 MFI. **b.** Antigen-specific T cell activation was assessed by measuring CD69 upregulation by flow cytometry. TPR Jurkat cells expressing TCR50, which recognizes the EBV LMP2 epitope (CLGGLLTMV) presented by HLA-A02:01, were used as a positive control. TCR^-^ TPR Jurkat cells served as a negative control. Stimulation with αCD3/αCD28 and PMA–ionomycin were included as assay controls. TPR Jurkats cells expressing TCR12, TCR13, TCR22 or TCR42 upregulated CD69 following co-culture with Ham014 APCs pulsed with EBV-derived, but not influenza-derived, peptides. **c.** UMAP visualization of EBV peptide–recognizing T cells, showing that virus-specific T cells are largely effector memory cells.

**Fig. S15.**
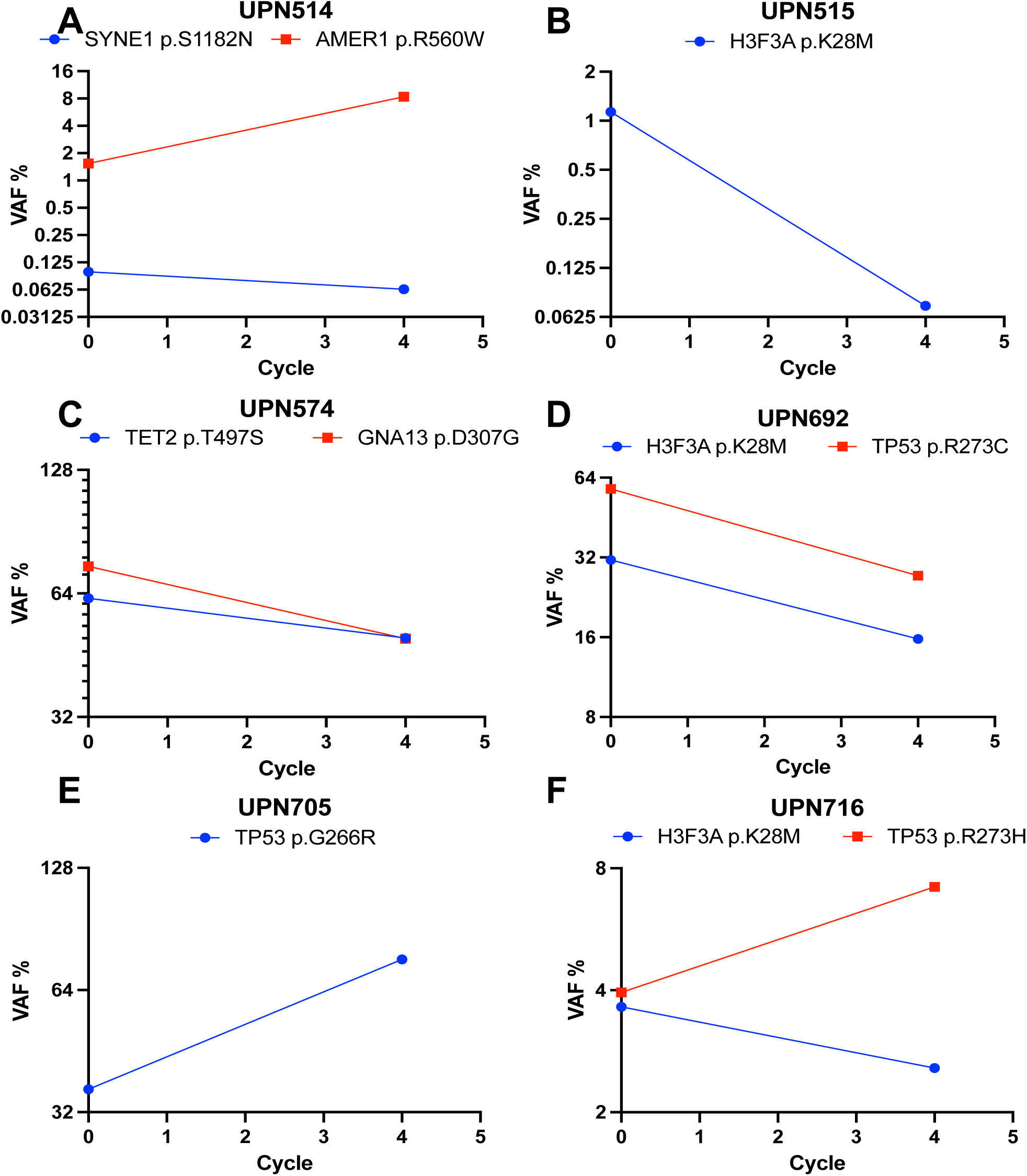
Longitudinal ctDNA analysis of patient CSF. Patient CSF samples from C1D0 (pre-treatment) and C4D0 (21 days into treatment) were submitted for targeted sequencing of known somatic mutations. Trends in variant allele fraction (VAF) were mostly consistent with radiographic course for all patients. A. UPN514 had a mild radiographic decrease in tumor size, but this was not seen until after Cycle 4. B. UPN515 showed a VAF decrease from ∼1%, matching a radiographic response. C,D. UPN574 and UPN694 did not have any radiographic response, but did show a shared decrease in VAF over two targets. Unfortunately, later sampling timepoints were not available. E. UPN705 did not respond to treatment and showed an increase in VAF. F. UPN716 showed a marked divergence in signal between two targets (H3F3A K27M decreased while TP53 R273H increased). This is consistent with a mixed radiographic response, and also supports the importance of multi-target measurement to capture tumor evolution during treatment response monitoring.

**Fig. S16.**
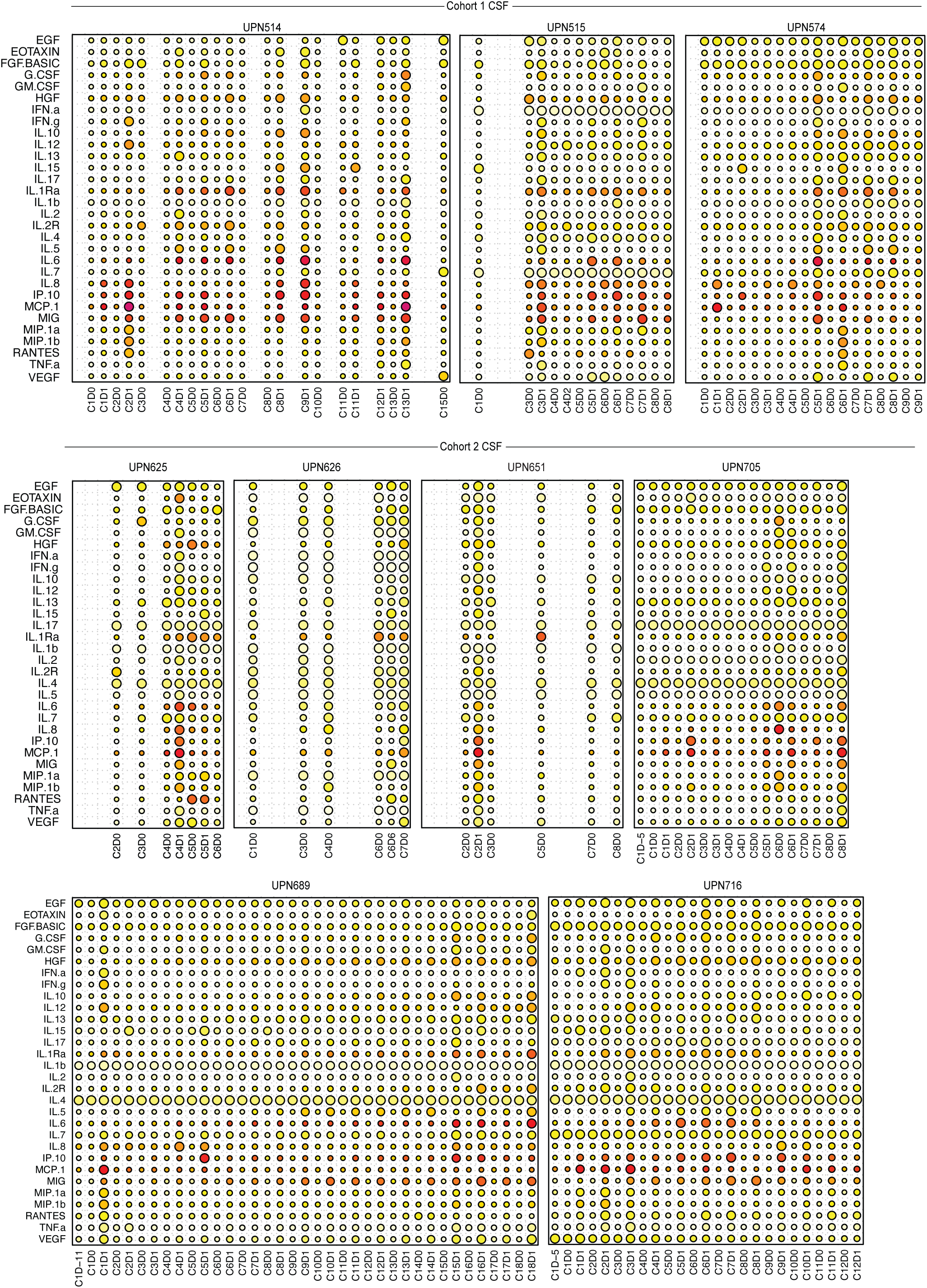
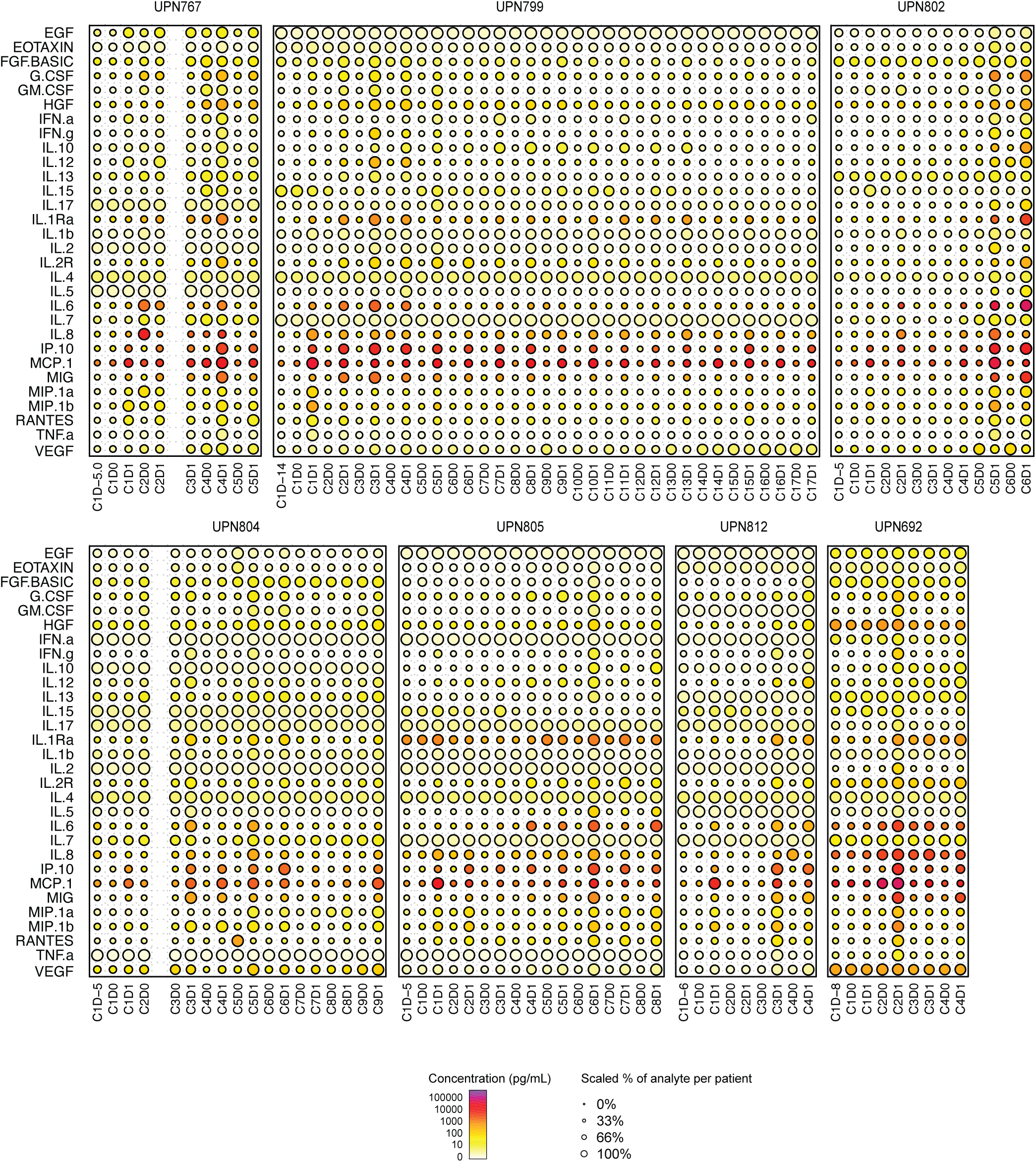
Cytokine concentrations in CSF are shown for at sampled timepoints for each patient during CAR-T therapy. The concentration of each cytokine (in pg/ml) at a given timepoint is denoted by the color of the corresponding dot in the relevant plot, with the color to concentration scale as shown (e.g.purple/red corresponds to the highest levels, white/yellow to the lowest). Additionally, the concentration relative to the maximum concentration observed for a given patient, compartment and analyte are shown by the size of the dots, as given (i.e. the maximum value of an analyte for a given patient and compartment, is represented by a dot corresponding to size 100% in the scale, while all other analytes are represent by an appropriately scaled dot, scaled by their percentage relevant to the observed maximum).

**Fig. S17.**
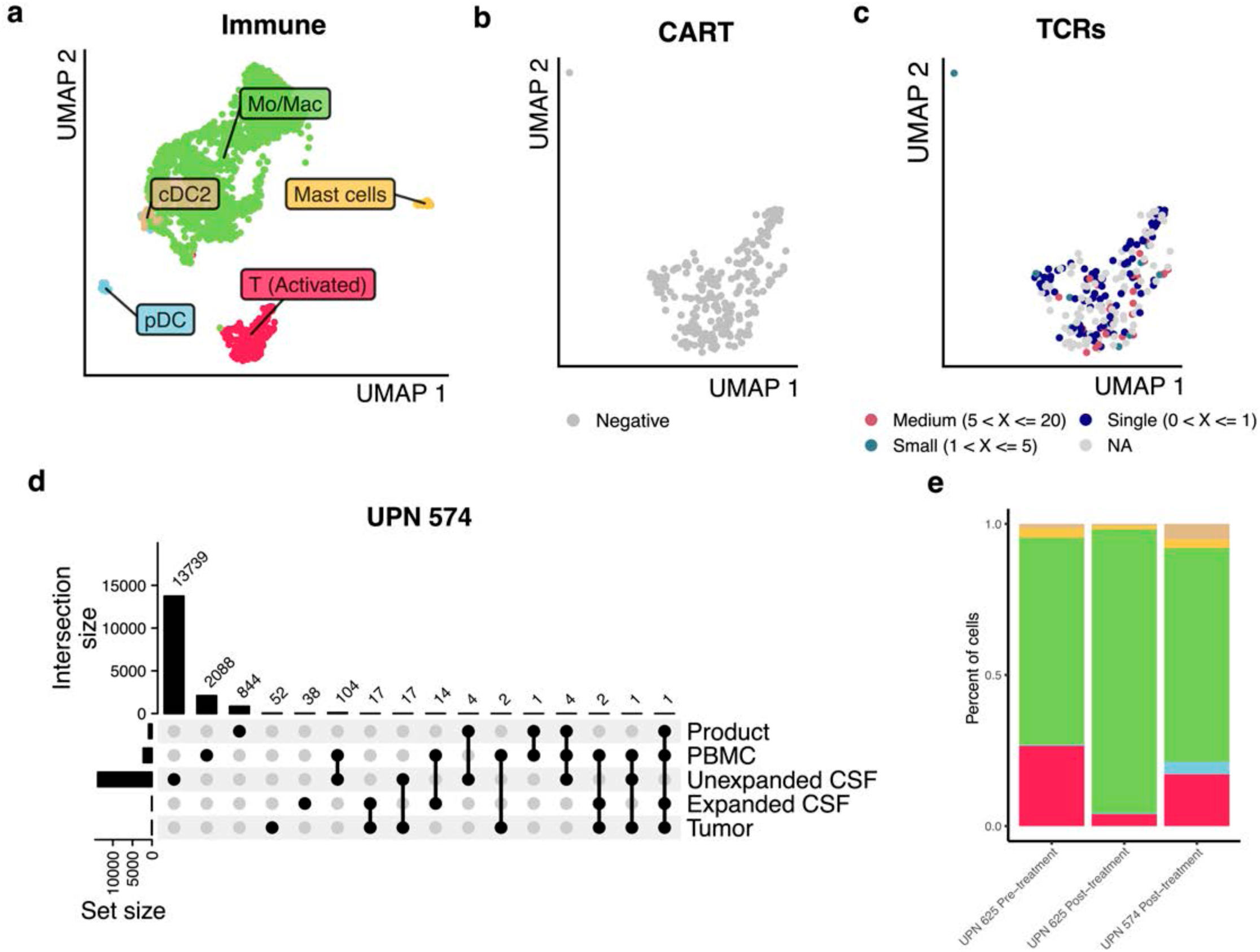
Characterization of immune cells in tumor. UMAP of A) 1,891 immune cells from tumor across two patients (UPNs 574 and 625), colored cell type, and 233 T cells colored by B) CAR positivity and C) TCR frequency. TCR data were only recovered from 625 pre- and 574 post-treatment tumors so all cells for 625 post-treatment tumors are colored gray. There were no CAR+ T cells observed in these tumor samples. D) Upset plot comparing all TCR clonotypes among tumor, CSF, PBMC, and product for UPN 574. CSF TCRs are separated by whether they have an expanded or unexpanded TCR frequency. 17 TCRs found in the tumor overlapped with expanded TCRs found in the CSF (of 72 total expanded TCRs in CSF); 17 TCRs found in the tumor overlapped with unexpanded TCRs found in the CSF (of 13,869 total unexpanded TCRs in CSF). E) Stacked bar plot of cell type proportions for each sequenced tumor sample.

**Fig. S18.**
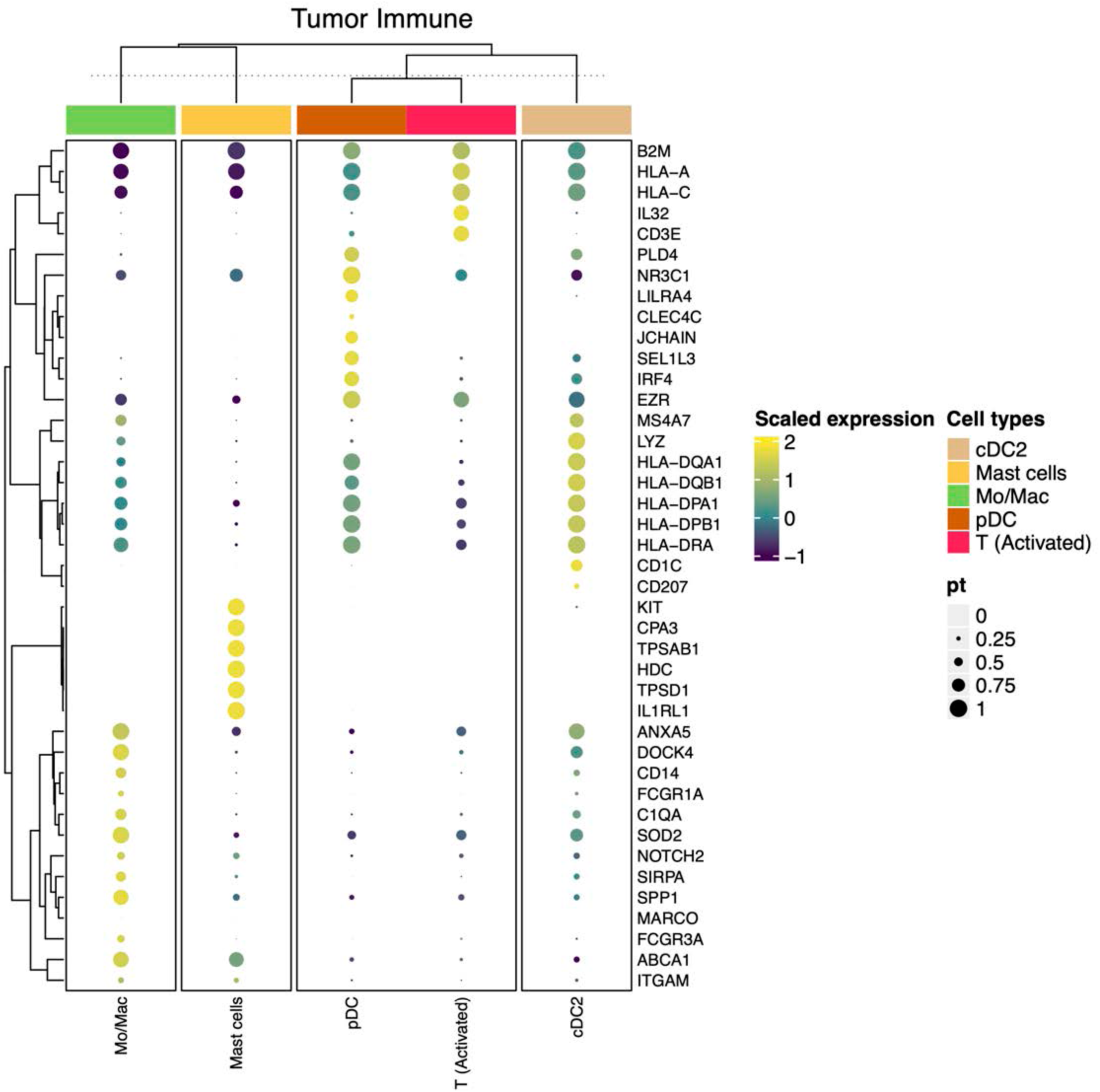
Dot plot heatmap of genes used in cell type annotations for immune cells in tumor. Top 5 markers were identified for each cell along with a curated set of canonical markers.

**Fig. S19.**
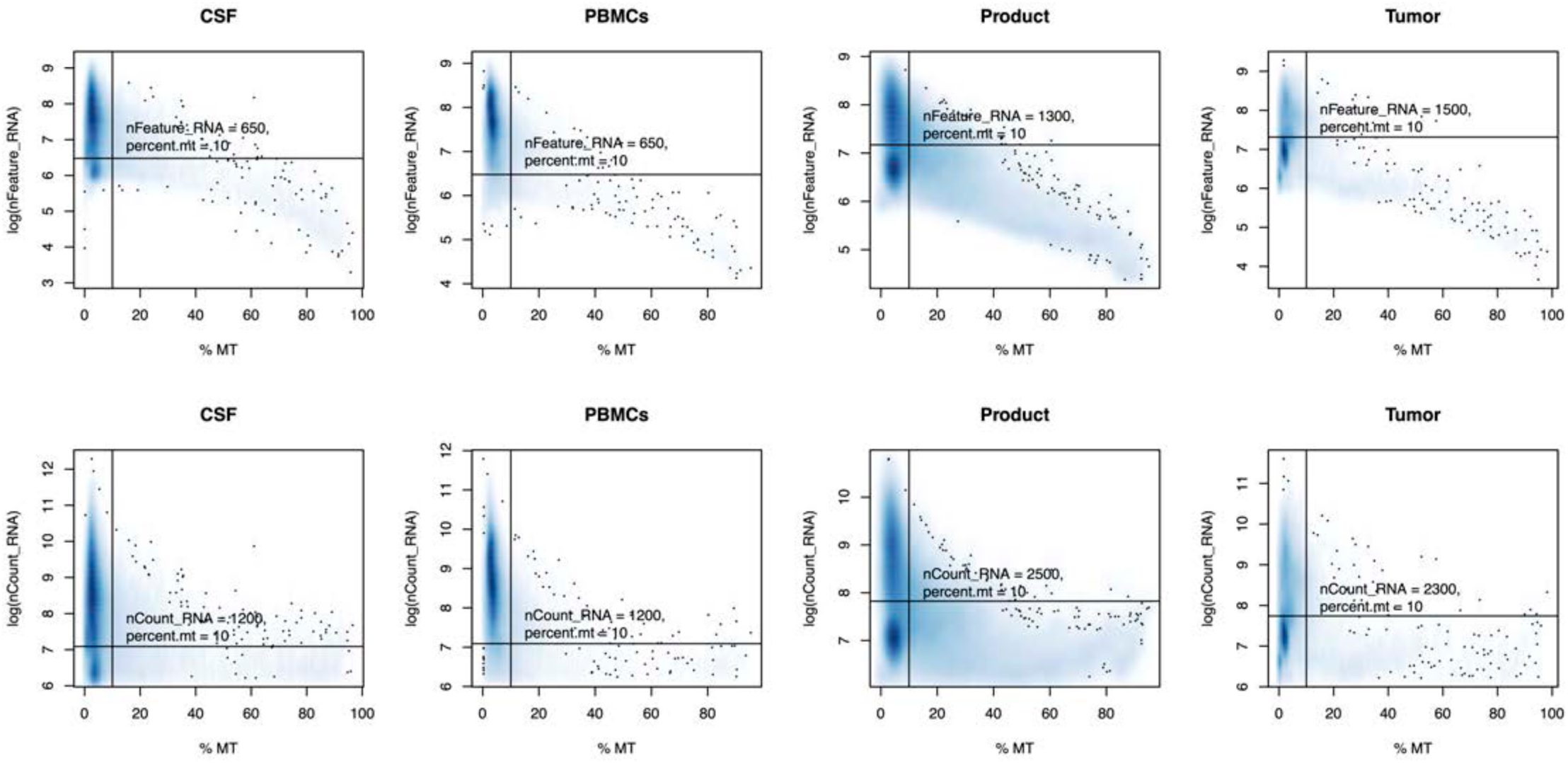
Filtering thresholds. Smoothscatter plots for each sample type showing the distribution of %mt and log(nFeature) (top) and %mt and log(nCount) (bottom). Vertical and horizontal lines represent filtering thresholds implemented on each sample type.

**Fig. S20.**
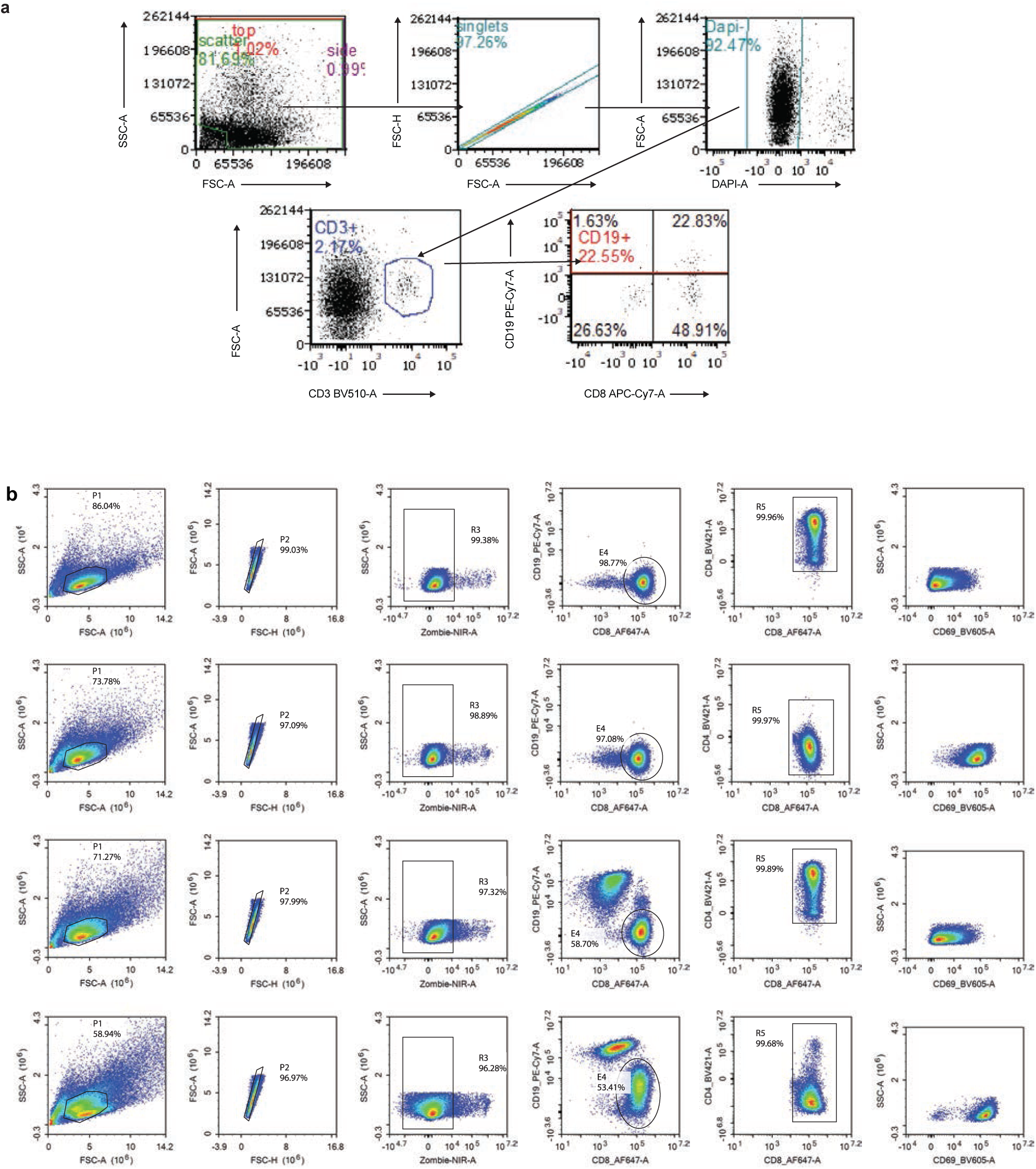
Gating strategies. **a.** Gating strategy for identifying CAR^+^ T cells by flow cytometry, using CD19t transduction tag. **b.** Flow cytometry gating strategy for the TPR Jurkat co-culture assay. Representative gating plots used to quantify activation of TPR Jurkat cells expressing TCR37 (UPN514), measured by median fluorescence intensity (MFI) of CD69. Conditions shown (top to bottom) include untreated cells, stimulation with PMA/ionomycin, and co-culture with Ham014^WT^ or Ham014^CAR^ cells.

